# Healthcare service user-reported quality of care in Malawi: a national multi-facility cross-sectional study

**DOI:** 10.1101/2025.09.10.25335306

**Authors:** Joseph H Collins, Wiktoria Tafesse, Precious Chitsulo, Timothy B Hallett, Eva Janoušková, Joseph Mfutso-Bengo, Emmanuel Mnjowe, Sakshi Mohan, Watipaso Mulwafu, Dominic Nkhoma, Bingling She, Mariana Suarez, Tim Colbourn

## Abstract

**Background:** Improving healthcare service user satisfaction is one of the three core objectives of the Malawian government’s health sector strategic plan. As such, robust understanding of service users’ perceptions of care quality is crucial to inform patient-centred services that can drive service improvements.

**Objective:** This study aimed to explore how service users in Malawi perceive the quality of the healthcare they receive across services and facilities and to investigate the association between individual and health service characteristics and perceived quality of care.

**Design:** A national multi-facility cross-sectional study using service user exit interview data.

**Setting(s):** 30 health facilities across 15 districts in Malawi.

**Participants:** 4,181 respondents surveyed after completing their visit and exiting healthcare facilities between January and May 2024.

**Methods:** Descriptive statistics of service user-reported quality of care and multinomial logistic regression analyses to determine the association between patient and healthcare characteristics and perceived care quality.

**Results:** Quality of care was reported as being high with 58% of respondents rating care as ‘Good’ and 35% as ‘Very good’, with some variation by ‘dimension’ of care (e.g. treatment availability). Positive or negative perceptions of care quality were associated with age, sex, education level, illness severity, previously seeking care, referral, facility type, non-governmental facility ownership, service area, access to medication and payment of fees.

**Conclusions:** Most patients in Malawi report receiving high-quality care, although a minority perceive the quality to be inadequate. Action to address gaps in service delivery, such as improving the availability of required medicines, could address the poor perceptions of quality held by the minority of interviewed service users.

**What is already known on this topic:**

- Service user-reported quality of care is an important indicator of quality healthcare delivery and key for responsive and patient-centred systems.
- Previous assessments from Malawi are largely focused on single services, facilities or groups.

**What this study adds:**

- This study reveals that, across a variety of facilities and services, most Malawians rate the quality of care they receive highly whilst a minority reported quality of care to be neither good nor bad, bad, or very bad.
- We identified that non-governmental facility ownership and factors related to how services are delivered (i.e. availability of prescribed medication, patient referral, service area) are strongly associated with perceived quality.

**How this study might affect research, practice or policy:**

- Several important factors associated with service user-reported quality, including availability of medication and referral systems, are amenable to policy action and could drive improvements in both quality and health. Additionally, we highlight the need for research into the factors influencing perceived quality in non-governmental facilities in Malawi.

## Introduction

Despite the concerted efforts of national governments during the Sustainable Development Goal (SDG) era, including those prompted by global initiatives such as the Lancet Global Health Commission on High Quality Health Systems (1) and World Health Organization coordinated Quality of Care Network (2), there remains substantial inter- and intranational variation in the quality of healthcare (3,4). Poor quality health services are a key barrier to progress in global public health, given the relationship between quality and health outcomes (5). This is particularly true in low- and middle-income countries (LMICs) such as Malawi, in which system-wide drivers of poor-quality care include limited availability of medical consumables, healthcare workers and functional infrastructure (6), driven by significant constraints on resources for health (7), compounded by a growing burden of communicable and non-communicable disease (8).

Given these challenges, empirical evaluation of service quality is needed to guide service delivery improvements. The components of healthcare quality, such as effectiveness, safety and people-centredness, can be assessed using objective measures, in which observed care is compared to pre-defined quality or clinical indicators, or subjective measures, where service users report their perceptions of the quality of care they have received (9,10). Commonly, subjective evaluation includes elicitation of service users’ satisfaction or experiences across several dimensions of care delivery via a survey instrument. High-quality care is responsive to the preferences, needs and values of service users (11), and service users are best placed to assess the quality of a product (11). Therefore, understanding service user-reported quality is vital, particularly given the relationship between perceived quality and healthcare seeking behaviour (11). Importantly, service user experience of healthcare has been found to have a positive association with clinical effectiveness and safety across disease areas and settings (12), suggesting service users’ perceptions can provide a valid measure of overall healthcare quality.

Malawi’s Ministry of Health (MoH) continues to demonstrate a clear commitment to the universal delivery of high-quality health services through the development of the Quality Management Directorate (QMD) in 2016 and operationalisation of the most recent Health Sector Strategic Plan (HSSP) III (2023-2030) (13). Key quality improvement initiatives include the implementation of the first national Quality Management Policy (2016-2022) (14) and a novel quality rating program that standardizes facility assessments and designates ‘Centers of Excellence’ (13). High-quality health systems must not only be reliable and adaptive (15), but they must also be trusted by users and responsive to their needs (16). Thus, the MoH has placed improving client satisfaction as central in moving towards Universal Health Coverage (UHC) (13).

However, it is difficult to draw conclusions related to how service users perceive the quality of the services they receive in Malawi from the existing empirical literature given the heterogeneity of included samples and health services. Most studies have explored user perceptions of quality in relation to one or two services or service areas (6,17–25) with a particular focus on reproductive and maternal health (6,17,18,22–25). Where quality of multiple services is explored, this is largely done within single facilities, which are mostly at the tertiary level of the health system (18,26). Studies focus almost exclusively on adolescent or adult patients (17–25), with few studies exploring self-reported QoC for children (6).

This study seeks to (i) explore how service users in Malawi perceive the quality of the health care they receive across a variety of healthcare services and facilities; (ii) investigate the association between key individual and health service characteristics, on the one hand, and perceived quality of care, on the other.

## Methods

### Study design

We report here an analysis of data collected as part of the *Thanzi La Mawa* (TLM) study - a cross-sectional and mixed methods study intended to improve understanding of healthcare delivery in Malawi and to inform Malawi’s first whole-health system mathematical model (the *Thanzi La Onse* model (TLO)) (27). The TLM study included facility audits, time-and-motion data collection, service user exit and follow-up surveys, and qualitative interviews of facility managers. The study design and data-collection processes have been previously described (28). Data analysed for this study was collected primarily through service user exit-surveys and we have used the STROBE guidelines on cross-sectional studies (29) to report these results (Supplementary File 1 – Table S1).

### Setting

The service user exit-survey component of the TLM study was conducted at 30 health facilities across 15 districts in Malawi namely Blantyre, Chiradzulu, Dedza, Karonga, Likoma, Lilongwe, Mzimba, Mzuzu, Neno, Nkhata Bay, Nsanje, Ntcheu, Rumphi, Thyolo and Zomba. Healthcare services are delivered in Malawi via a three-level system consisting of primary, secondary, and tertiary levels, with a referral system operating between them (13). At the primary level, services are delivered to the community via health posts, dispensaries, health centres and community hospitals. At the secondary level, inpatient and outpatient services are delivered via Government district hospitals and other non-state hospitals of similar capacity. Tertiary level services are provided by four central hospitals and one mental hospital, which act as referral centres for secondary hospitals. Whilst the health system consists of a mix of public, private-for-profit and private not-for-profit service providers, the public sector, which delivers services for free at the point of use, provides the majority of healthcare (6). The private sector provides the remainder of services, with the Christian Health Association of Malawi (CHAM), a faith-based private-not-for-profit organisation, being the largest private provider (6). The Government has a Memorandum of Understanding with CHAM, under which neither party constructs a facility within a 7 km radius of the other. In turn, Service Level Agreements between the Government and CHAM providers reimburse CHAM for part of the service costs, thereby enabling users to access contracted services free of charge.

### Participants and data collection

Data was collected between January and May 2024 at 30 health facilities including all four central (tertiary) hospitals (Queen Elizabeth, Zomba, Kamuzu, Mzuzu) and Zomba mental hospital. To ensure a representative sample 25 facilities were selected visa stratified random sampling. Facilities were selected using a random number generator in Microsoft Excel from the National Master Health Facility Register following stratification by facility type (health centre, community hospital, secondary hospital), patient volume (relatively lower or higher), ownership (MOH or CHAM), and location (urban or rural). A full list of included facilities is available in Supplementary File 1 – Table S2.

Exit-survey data was collected by five teams of enumerators across enrolled facilities using KoboToolbox (30), a secure data-collection software allowing for offline data capture and data validation, following enumerator training and initial data-collection tool piloting in January 2024. Enumerators randomly selected potential participants from adults exiting the facility after being discharged or accompanying someone who was discharged. To survey a representative random sample of patients, participant selection was based on daily patient load (estimated via facility registers or waiting room attendance). Given enumerators aimed to survey 15 patients per day, every X^th^ patient was interviewed to reach this number of surveys (e.g. if 60 people were waiting for treatment, every 4^th^ person was interviewed). The survey was formulated and coded in English and translated by enumerators to Chichewa and Tumbuka as required during data collection. Informed consent was obtained prior to participation. Enumerators were trained to ensure respondents were out of earshot of facility workers and to encourage respondents to speak freely. There were no restrictions on eligibility for participation.

The service user exit-survey, available in Supplementary File 2, consisted of possible 68 questions across seven areas; patient details (11 questions), facility visit (14 questions), patients’ access to medicines (2 questions), patients’ persistence in care-seeking (3 questions), costs and opportunity costs of attending facility (11 questions), household assets (17 questions) and perceptions of quality of care (10 questions). In total, data was collected from 4,181 respondents across all included facilities

Following data collection, data was extracted from KoboToolbox to Microsoft Excel (Office 365, Version 16.94), anonymised and imported into Stata/SE 18.0 (Statacorp) for data cleaning and analysis. Supplementary File 3 is the Stata.do file describing how the data was cleaned.

### Statistical analysis

Characteristics of the sample, the included facilities, and the service user-reported quality of care have been described first with descriptive statistics. The association between variables and the primary outcome, overall service user-reported quality of care for the facility visit, was explored via multinomial logistic regression. The primary outcome of this analysis was a categorical variable derived from the Likert scale survey statement assessing ‘overall’ quality of the participants visit, with three responses: ‘Very good’, ‘Good’, ‘Neutral -Very bad’. Using those who reported that care was ‘Good’ as the reference category allowed for focus of analytical power on the extremes of responses and to account for the small number of respondents who rated overall quality as ‘Bad’ or ‘Very bad’.

Covariates selection was informed by the existing literature describing determinants of service user-reported quality of care or satisfaction reported in studies from Malawi (6,17– 20,22–25) and a systematic review evaluating determinants of service user-reported satisfaction with care delivered across a given health system (31). All covariates included are described in Table 1. Where covariates are categorical or binary variables derived from survey questions, Table S3 - Supplementary File 1 describes the relevant derivation. Missing data was observed in six variables, including service user-reported quality of care (1.6%), and ranged from 0.4% - 10.2% of total entries. To assess if missing data in the primary outcome was clustered by facility, we conducted a Chi-square test of independence between the missingness indicator and facility.

**Table 1.**
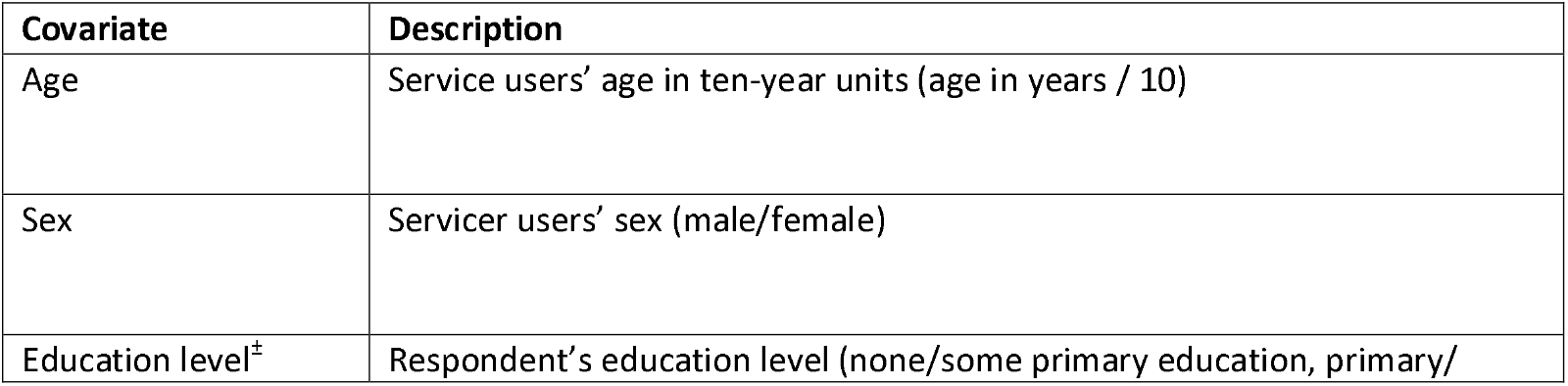
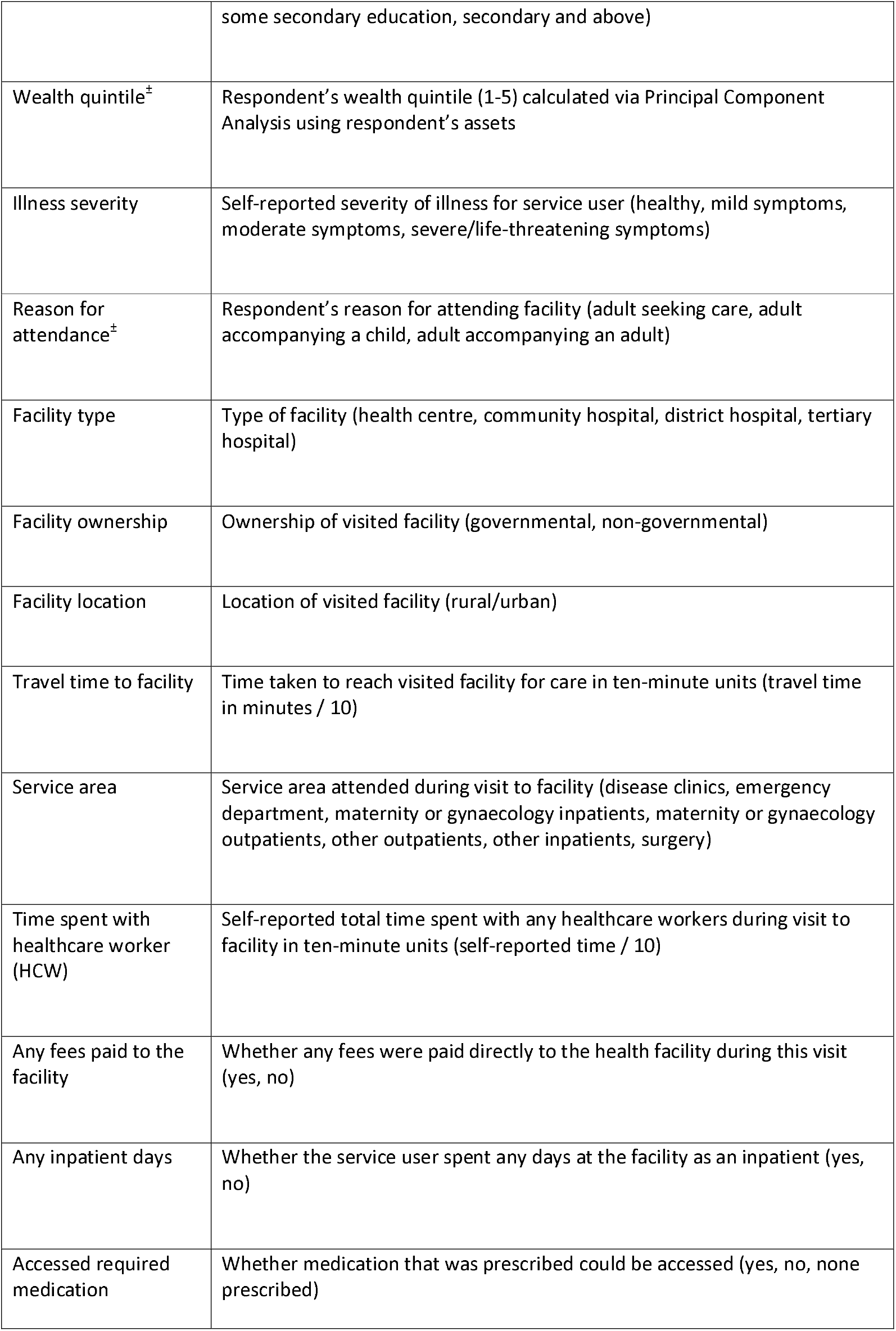
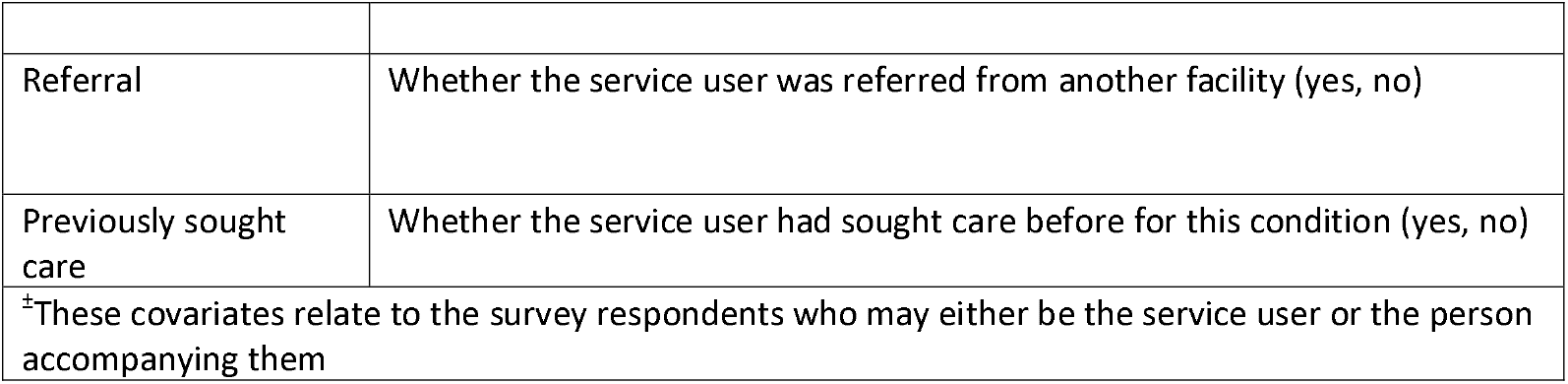
Description of covariates

We developed three multivariable models, each incrementally incorporating additional variables to systematically address confounding. Individuals with missing outcome or covariate data were excluded from regression models. Model 1 included variables to address confounding effects driven by individual service user sociodemographic and illness characteristics (age, sex, education level, wealth quintile, illness severity, previously sought care, referral). Model 2 included all variables in Model 1 plus facility characteristics (type, ownership, location) and service characteristics (service area, any inpatient days). Finally, in Model 3, we add policy-relevant variables closely related to service delivery (travel time to facility, time spent with a healthcare worker (HCW), access to required medication, any fee paid to facility). Collinearity between covariates was assessed via Variance Influence Factors. Model standard errors were estimated using the variance-covariance estimator to account for intra-group correlation within facilities. For all tests a p-value lower than 0.05 (95% confidence interval not crossing 1, no effect) was considered to indicate statistical significance.

We conducted the same analyses with outcome variables derived from each of the Likert survey questions assessing perceptions of quality relating to healthcare worker consultation, respect, waiting time and treatment availability to provide a more comprehensive view of quality perception. The Stata.do analysis file is available as Supplementary File 4. Data required to reproduce the study’s results is available upon request.

### Ethical approval

Ethical approval for the TLM study was provided by Kamuzu University of Health Sciences Ethics Committee (COMREC) on 15/11/2023. Kamuzu University of Health Sciences carried out the data collection and are the designated sponsor.

## Results

### Sample characteristics

The sociodemographic and illness characteristics of the sample are summarised in Table 2, alongside key characteristics of the facilities at which they were interviewed. The mean age of respondents in the total sample was 25.7 years (Standard Deviation (SD) 18.5). More of the total sample were women (62.1%, n=2,957) with nearly half of participants having received less than a complete primary education (41.9%, n=1750) and the highest percentage of participants were in the highest wealth quintile (25.1%, n=1,051). Participants were most likely to report severity of illness as mild (39.9%, n=1,667) which was reflected in the services participants accessed with the largest proportion attending ‘Other clinics/outpatients’ care (64.5%, n=2,696). Respondents were evenly split between being interviewed leaving a community hospital (31.8%, n=1,330), tertiary hospital (29.7%, n=1,241) or health centre (27.5%, n=1,149) with fewer interviewed leaving a district hospital (Table 2). 63.8% (n=2,667) of the service user sample received care in government facilities and 69.1% (n=2,888) in urban settings.

**Table 2.** Sample and facility characteristics

|  | **Total**<br>**sample** |
| --- | --- |
| *(N)* | *4,181* |
| **Age (Years)** |  |
| *Mean, SD* | 25.7 (18.5) |
| **Sex:** |  |
| *Male* | 2,597 (62.1%) |
| *Female* | 1,584 (37.9%) |
| **Education level:** |  |
| *None / Some primary* | 1,750 (41.9%) |
| *Primary / Some secondary* | 1,635 (39.1%) |
| *Secondary and above* | 778 (18.6%) |
| *<u>Missing</u>* | 18 (0.4%) |
| **Wealth quintile:** |  |
| *1st (highest)* | 1,051 (25.1%) |
| *2nd* | 654 (15.6%) |
| *3rd* | 821 (19.6%) |
| *4th* | 823 (19.7%) |
| *5th* | 832 (19.9%) |
| **Illness severity:** |  |
| *Healthy* | 859 (20.5%) |
| *Mild symptoms* | 1,667 (39.9%) |
| *Moderate symptoms* | 1,385 (33.1%) |
| *Severe/Life-threatening symptoms* | 270 (6.5%) |
| **Reason for attendance:** |  |
| *Adult seeking care* | 2,527 (60.4%) |
| *Adult accompanying another adult* | 272 (6.5%) |
| *Adult accompanying a child* | 1,382 (33.1%) |
| **Previously sought care** |  |
| *Yes* | 1,888 (45.2%) |
| *No* | 2,273 (54.4%) |
| *<u>Missing</u>* | 20 (0.5%) |
| **Referral** |  |
| *Yes* | 652 (15.6%) |
| *No* | 3,515 (84.0%) |
| *<u>Missing</u>* | 16 (0.4%) |
| **Facility type:** |  |
| *Health centre* | 1,149 (27.5%) |
| *Community hospital* | 1,330 (31.8%) |
| *District hospital* | 461 (11.0%) |
| *Tertiary hospital* | 1,241 (29.7%) |
| **Facility ownership:** |  |
| *Governmental* | 2,667 (63.8%) |
| *Non-governmental* | 1,514 (36.2%) |
| **Facility location:** |  |
| *Urban* | 2,888 (69.1%) |
| *Rural* | 1,293 (30.9%) |
| **Service area:** |  |
| *Disease clinics* | 514 (12.3%) |
| *Emergency department* | 59 (1.4%) |
| *Maternity/Gynaecology inpatient* | 225 (5.4%) |
| *Maternity/Gynaecology outpatient* | 398 (9.5%) |
| *Other clinics/outpatients* | 2,696 (64.5%) |
| *Other inpatient wards* | 237 (5.7%) |
| *Surgery* | 52 (1.2%) |
| **Any inpatient days** |  |
| *Yes* | 984 (23.5%) |
| *No* | 3,197 (76.5%) |
| **Travel time to facility (Minutes)** |  |
| *Mean, SD* | 65.8 (49.5) |
| *Missing (n, %)* | 61 (1.46%) |
| **Time spent with HCW (Minutes)** |  |
| *Mean, SD* | 23.1 (40.7) |
| *Missing (n, %)* | 428 (10.2%) |
| **Access to required medication** |  |
| *Yes* | 3,528 (84.4 %) |
| *No* | 348 (8.3%) |
| *None prescribed* | 305 (7.3%) |
| **Any fees paid to the facility** |  |
| *Yes* | 859 (20.6%) |
| *No* | 3,322 (79.5%) |

### Service user-reported quality of care

Service user-reported quality of care is summarised in Table 3. Most respondents rated the overall quality of the care they received as either ‘Very good’ (34.7%, n=1,450) or ‘Good’ (57.6%, n=2,406). Over 92% of participants responded that they felt the care they received was appropriate. When considering quality across specific dimensions there was greater variation. The greatest number of participants reported care was ‘Bad’ or ‘Very bad’ when asked to rate quality in relation to waiting times (‘Very bad’ 2.5% (n=104), ‘Bad’ 7.5% (n=313)), followed by treatment availability (‘Very bad’ – 0.7% (n=30), ‘Bad’ 3.6% (n=148). When combining ‘Neither good nor bad’, ‘Bad’ and ‘Very bad’ responses relating to overall quality of care for the primary outcome of the regression analysis, 6.2% (n=259) of the sample reported the overall quality of care was at this level.

**Table 3.** Service user-reported quality of care in Malawi

|  | **Total**<br>**Sample**<br>**(n=4,181)** |
| --- | --- |
| **Health worker consultation:** |  |
| *Very good* | 1,651 (39.49%) |
| *Good* | 2,380 (56.92%) |
| *Neither good nor bad* | 71 (1.70%) |
| *Bad* | 29 (0.69%) |
| *Very bad* | 6 (0.14%) |
| <u>*Missing*</u> | 44 (1.05%) |
| **Treated with respect:** |  |
| *Very good* | 1,727 (41.31%) |
| *Good* | 2,300 (55.01%) |
| *Neither good nor bad* | 88 (2.10%) |
| *Bad* | 18 (0.43%) |
| *Very bad* | 6 (0.14%) |
| <u>*Missing*</u> | 42 (1.00%) |
| **Waiting time:** |  |
| *Very good* | 1,282 (30.66%) |
| *Good* | 2,180 (52.14%) |
| *Neither good nor bad* | 248 (5.93%) |
| *Bad* | 313 (7.49%) |
| *Very bad* | 104 (2.49%) |
| <u>*Missing*</u> | 54 (1.29%) |
| **Treatment available:** |  |
| *Very good* | 1,646 (40.12%) |
| *Good* | 1,982 (48.31%) |
| *Neither good nor bad* | 297 (7.24%) |
| *Bad* | 148 (3.61%) |
| *Very bad* | 30 (0.73%) |
| <u>*Missing*</u> | 78 (1.87%) |
| **Overall:** |  |
| *Very good* | 1,450 (34.68%) |
| *Good* | 2,406 (57.55%) |
| *Neither good nor bad* | 196 (4.69%) |
| *Bad* | 52 (1.24%) |
| *Very bad* | 11 (0.26%) |
| <u>*Missing*</u> | 66 (1.58%) |
| **Appropriate care received:** |  |
| *Yes* | 3,867 (92.49%) |
| *No* | 314 (7.51%) |

### Bivariate regression analysis

Table 4 reports the independent association between the variables and the primary outcome. The Chi-square test of independence revealed no significant association between the missingness of the primary outcome and facility (p = 0.498). Age, education level, illness severity, reason for attendance, facility type, facility ownership, facility location, service area, any fees paid to the facility, any inpatient days, access to required medications, referral, previously seeking care were all associated with respondents having either a more negative or very positive perception of quality of care compared to perceiving care as ‘good’. Wealth quintile and time spent with HCW were only associated with respondents having a more positive perception of quality of care whilst travel time to facility was only associated with a more negative perception of quality of care.

**Table 4.** Bivariate analysis of individual and health service factors and service user-reported quality of care

| VARIABLES | Service user-reported Quality of Care |  |  |
| --- | --- | --- | --- |
|  | Unadjusted Relative Risk Ratio (RRR) (95% CI) |  |  |
|  | (Reference outcome - Good) |  |  |
|  | Neutral – Very bad | Very Good | Observations |
| **Age** (10-year units) | 1.13***<br>(1.06, 1.21) | 1.04**<br>(1.00, 1.08) | 4,115 |
| **Sex** (male) | 1.14<br>(0.88, 1.48) | 0.90<br>(0.79, 1.03) | 4,115 |
| **Education level** (*Ref – None to some primary*) |  |  | 4,098 |
| *Primary to some secondary* | 0.66***<br>(0.50, 0.88) | 1.17**<br>(1.01, 1.36) |  |
| *Secondary and above* | 0.76<br>(0.52, 1.11) | 1.90***<br>(1.59, 2.27) |  |
| **Wealth quintile** (*Ref – First quintile (highest)*) |  |  | 4,155 |
| *Second quintile* | 1.00<br>(0.67, 1.48) | 1.11<br>(0.89, 1.37) |  |
| *Third quintile* | 0.91<br>(0.62, 1.33) | 1.17<br>(0.96, 1.43) |  |
| *Fourth quintile* | 0.98<br>(0.67, 1.43) | 1.35***<br>(1.11, 1.65) |  |
| *Fifth quintile* | 0.99<br>(0.67, 1.46) | 1.81***<br>(1.49, 2.02) |  |
| **Illness severity** (*Ref – Healthy*) |  |  | 4,155 |
| *Mild symptoms* | 1.12<br>(0.72, 1.74) | 0.27***<br>(0.23, 0.33) |  |
| *Moderate symptoms* | 1.17<br>(0.75, 1.83) | 0.23***<br>(0.19, 0.27) |  |
| *Severe/life-threatening symptoms* | 2.60***<br>(1.50, 4.49) | 0.37***<br>(0.27, 0.50) |  |
| **Reason for attendance** (*Ref – Adult seeking care*) |  |  | 4,115 |
| *Adult accompanying adult* | 0.76<br>(0.43, 1.35) | 1.04<br>(0.90, 1.35) |  |
| *Adult accompanying child* | 0.71**<br>(0.54, 0.95) | 0.70***<br>(0.62, 0.73) |  |
| **Previously sought care** (yes) | 1.82***<br>(1.40, 2.38) | 0.56***<br>(0.49, 0.64) | 4,097 |
| **Referral** (yes) | 2.88***<br>(2.10, 3.94) | 2.76***<br>(2.30, 3.30) | 4,099 |
| **Facility type** (*Ref – Health centre*) |  |  | 4,115 |
| *Community hospital* | 0.42**<br>(0.28, 0.61) | 1.48***<br>(1.25, 1.76) |  |
| *District hospital* | 0.47***<br>(0.29, 0.76) | 0.42***<br>(0.31, 0.56) |  |
| *Tertiary hospital* | 1.43**<br>(1.06, 1.93) | 2.20***<br>(1.84, 2.62) |  |
| **Facility ownership** (non-governmental) | 0.69**<br>(0.51, 0.92) | 1.36***<br>(1.91, 1.56) | 4,115 |
| **Facility location** (rural) | 0.64***<br>(0.48, 0.85) | 0.50***<br>(0.43, 0.59) | 4,115 |
| **Service area** (*Ref – Other clinics / outpatients*) |  |  | 4,115 |
| *Disease clinics* | 1.70**<br>(1.19, 2.44) | 2.12***<br>(1.73, 2.60) |  |
| *Emergency department* | 2.20<br>(0.83, 5.87) | 0.92***<br>(1.89, 5.67) |  |
| *Maternity/Gynaecology inpatient* | 1.32 | 9.30*** |  |
|  | (0.56, 3.14) | (6.63, 13.02) |  |
| *Maternity/Gynaecology outpatient* | 0.80 | 2.87*** |  |
|  | (0.45, 1.40) | (2.30, 3.57) |  |
| *Other inpatient wards* | 0.74 | 2.04*** |  |
|  | (0.37, 1.48) | (1.55, 2.70) |  |
| *Surgery* | 6.23*** | 6.00*** |  |
|  | (2.55, 15.25) | (3.12, 11.53) |  |
| **Any inpatient days (yes)** | 0.58*** | 2.51*** | 4,115 |
|  | (0.39, 0.87) | (2.16, 2.91) |  |
| **Travel time to facility (ten minutes)** | 1.04*** | 0.99 | 4,057 |
|  | (1.01, 1.06) | (0.98, 1.01) |  |
| **Time spent with HCW (ten minutes)** | 0.99 | 1.17*** | 3,693 |
|  | (0.92, 1.07) | (1.14, 1.20) |  |
| **Accessed required medication (Ref – Yes)** |  |  | 4,115 |
| *No* | 7.58*** | 0.12*** |  |
|  | (5.70, 10.08) | (0.07, 0.20) |  |
| *None prescribed* | 0.66 | 1.41** |  |
|  | (0.30, 1.44) | (1.11, 1.79) |  |
| **Any fees paid to the facility (Yes)** | 0.28*** | 0.78*** | 4,115 |
|  | (0.18, 0.45) | (0.66, 0.92) |  |

CI in parentheses \*\*\* p<0.01, \*\* p<0.05

### Multivariable regression analysis

Table 5 reports the adjusted associations between covariates and the primary outcome across the three models. We excluded the variable describing reason for attendance from multivariable modelling due to collinearity with age detected via Variance Influence Factors. Where variables are categorical, we provide the level associated with the primary outcome (compared to the reference category) in parentheses.

**Table 5.**
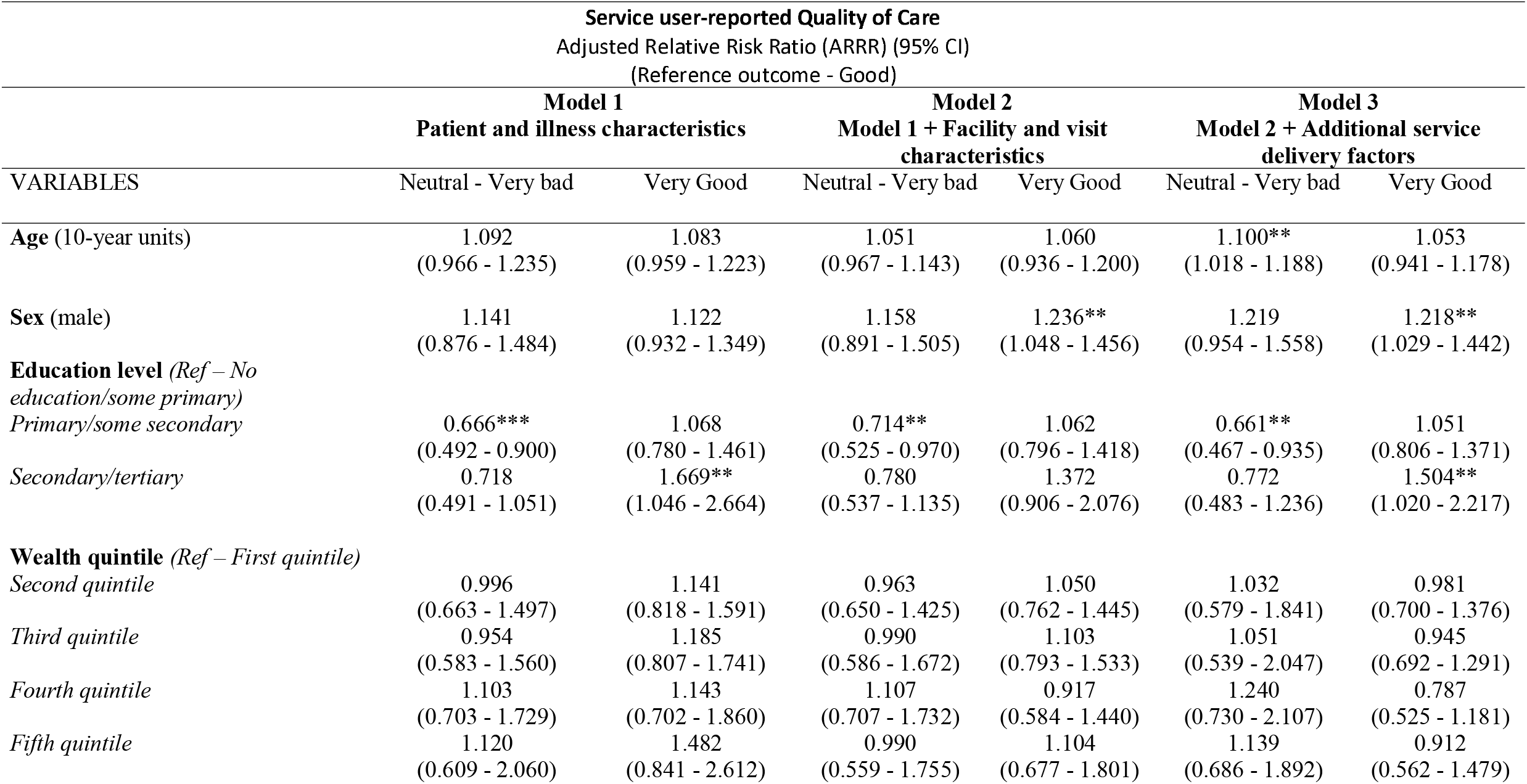
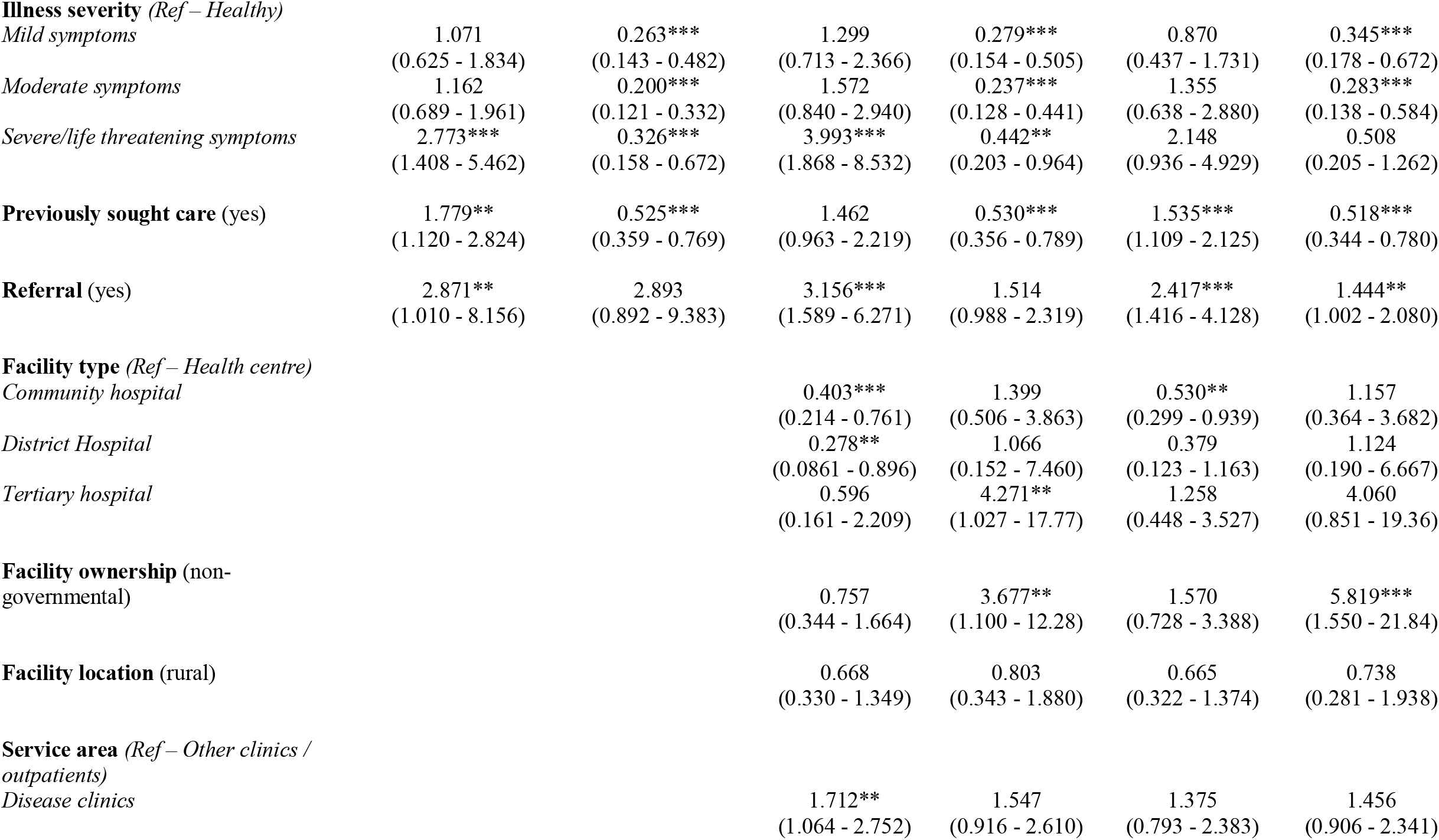
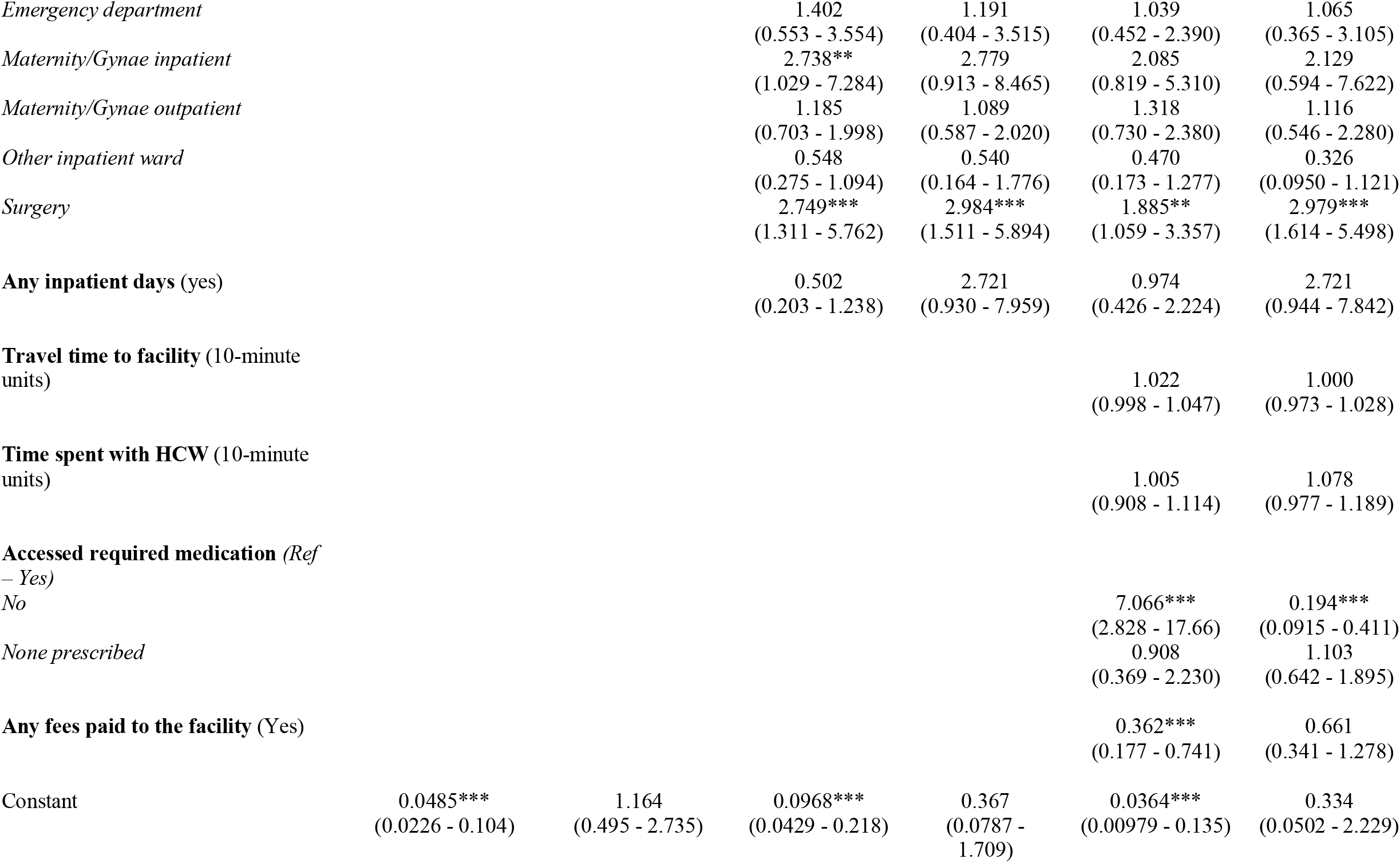
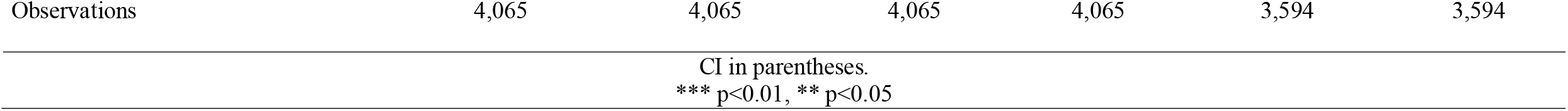
Multivariable analysis of individual and health service factors and service user-reported quality of care

In Model 1, age, sex and wealth quintile were not found to be associated with perceptions of care quality. Illness severity, having previously sought care, and referral were associated with respondents having a more negative perception of quality of care. Importantly, participants who were referred from another facility were nearly 3 times more likely to perceive care negatively (Adjusted Relative Risk Ratio (ARRR) 2.87, 95% Confidence Interval (CI) [1.01, 8.16]). Compared to participants who had completed none or some primary education, those who had completed primary/some secondary education were less likely to perceive care negatively. Participants with secondary/tertiary education were associated with having a more positive perception of quality of care.

Within Model 2, all facility and visit covariates, except facility location and any inpatient days, were associated with the primary outcome. Compared to those interviewed at health centres, individuals interviewed at tertiary hospitals were over 4 times more likely to perceive care positively (ARRR 4.27 (95% CI [1.04, 17.77])), whilst those at non-governmental facilities, compared to governmental facilities, were 3.7 times more likely (ARRR 3.68 (95% CI [1.10, 12.28])) to do so. Male sex was now found to be significantly associated with positive perceptions of care quality. Only primary/some secondary education level was significantly associated with less negative perceptions. While those who had previously sought care remained less likely to report very positive perceptions, the association with negative perceptions was no longer statistically significant.

Model 3 represents the final adjusted model including all covariates. In this model wealth quintile, facility location, any inpatient days, travel time to the facility and time spent with HCW were not associated with the primary outcome. Sex, education level (secondary/tertiary vs no education/some primary), illness severity (mild symptoms, moderate symptoms vs healthy), having previously sought care, referral, non-governmental facility ownership and service area (surgery vs. other outpatients) were associated with respondents having a more positive perception of care quality. Service users who received care in a non-governmental facility were nearly six-times more likely to perceive quality of care positively (ARRR 5.82 (95% CI [1.55, 21.84])). Age, education level (primary/some secondary vs no education/some primary), having previously sought care, referral, facility type (community hospital vs. health centre), access to required medication (no vs. yes) and paying any fees to the facility were found to be associated with respondents having a negative perception of care quality. Service-users who could not receive prescribed medication were over seven-times (ARRR 7.07 (95% CI [2.83, 17.66]) more likely to have a negative perception of care quality compared to those who could. Similarly, service users who could not access prescribed medication were much less likely to perceived care as being very good or better (ARRR 0.19 (95% CI 0.09, 0.412)).

Regression analyses exploring the relationships between covariates and perceptions of quality relating to healthcare worker consultation, respect, waiting time and treatment availability are presented in Tables S4-S7 (Supplementary File 1). In all models we observed that sex, illness severity, previously seeking care, referral, facility ownership, service area, access to required medication and any fees paid to the facility were significantly associated with perceptions of care quality with variation is the size of the effect or associated category.

We observed frequent differences between these models and Model 3 (Table 5) when examining associations with patient and illness characteristics. For example, age was not significantly associated with the perceptions of care quality in any of the other models, education level was only associated perceptions of quality relating to waiting time, and wealth was found to be associated with perceptions of quality relating to HCW consultation and treatment availability.

## Discussion

Empirical evaluation of how service users, the consumers of healthcare services, perceive the quality of the care they receive is central to improving system performance and health outcomes (4). Our study explores how over 4,000 service users in Malawi rate quality of care at 30 facilities across the country and the associations between key individual-level and health service determinants and perceptions of quality. We found that quality of care was perceived as being high across all evaluated dimension and identified several important determinants of perceived quality with large effect sizes related to facility ownership and service delivery factors such as the availability of prescribed medication and fees paid to the facility.

Our findings contribute to the existing academic literature from Malawi which describes both moderate to significant dissatisfaction with the quality of primary care services (20,21), specific facilities (26) or interpersonal care delivered during labour (22), alongside evidence of high or very high perceptions of health service quality (18,19,23–25,32). Whilst variation in these prior results is likely due to the breadth of methods applied, services evaluated and timing of evaluations, our study offers a contemporary, national, multi-service and multi-facility evaluation in which over ninety percent of respondents reported that the overall quality of care they, or those they accompanied, received was ‘Good’ or ‘Very good’. Comparatively, nationally representative evidence from Malawi, in which providers adherence to, or knowledge of clinical guidelines, was assessed, suggests quality of care across many services is low (6,33) despite substantial policy focus on quality improvement. Whilst higher levels of dissatisfaction with waiting times and the availability of treatment were observed in our study, likely associated with historic constraints on resources for healthcare impacting the availability of essential medical consumables (34,35) and trained healthcare professionals (13), we note the disparity between objective measures and subjective user-reported assessment of quality. This is likely driven by ‘consumers’’ expectations of healthcare (36,37). Service users across several LMICs were found to assess poor quality care as being good or better, when accounting for low technical awareness of correct treatment, suggesting differences are driven by low expectations of care (36). Importantly, service users with higher expectations are found to be more “active” in seeking and receiving higher quality health services (38,39) highlighting potential to improve health outcomes through increasing expectations.

Additionally, we identified a wide range of patient and healthcare characteristics that are associated with perceived quality in this population through regression analyses. At the individual level, several sociodemographic characteristics and self-perceived illness severity were found to be associated with quality, in line with the existing literature from beyond Malawi (31). Importantly, we found that the inability for service users to access prescribed treatment, compared to those who could, was the single largest predictor of rating care as negatively. Evidence suggests the availability of key medicines across the Malawian health system varies substantially by system level, facility type and ownership (34,35) with Mohan et al. identifying a mean availability for a subset of key medicines of just over fifty percent across 940 health facilities (34). Given our results, and that limited availability of medicines can impact health outcomes and drive out-of-pocket costs (40), concerted focus on policies to improve consumable availability, such as improvements to both access and management of drugs in lower level facilities (34), would likely drive improvements in perceptions of care quality and health.

Beyond medication availability, we found participants were nearly six-times more likely to rate care as ‘Very good’ compared to ‘Good’ if attending a non-governmental facility. Higher levels of patient satisfaction for those receiving care at Christian faith-based providers in Africa has been well documented (41,42) aligning with our own findings as CHAM facilities constituted most non-governmental facilities in the sample (38% of all facilities included, 85% of non-governmental facilities). This effect was even more pronounced in regression analyses in which perceived quality relating to waiting times was the primary outcome (Supplementary File 1, Table S6), suggesting that reduced waiting times in non-governmental facilities may partially drive more positive perceptions of quality. Additional reasons for increased satisfaction with care delivered by faith-based providers are unclear but may be due to delivery of more compassionate interpersonal care and the relationship between religion and organisational culture (41). However, evidence from Malawi suggests that for sexual and reproductive healthcare, service users at CHAM facilities are less likely to receive services related to prevention and investigation of sexually transmitted infections or promotion of condom use (43) suggesting despite high levels of satisfaction there are important shortcomings in service availability.

Whilst our study provides insights into service user-reported quality of care in Malawi, there are some limitations. While data collectors provided all participants with clear information about the study and independence between the research team and facility management, it is possible participants may have overestimated quality for fear of repercussions associated with lower ratings due to being interviewed near facility exit. Additionally, those who opted to undertake the exit interview may have some important differences compared to the wider care-seeking population as over two thirds of our sample were receiving outpatient care and because data from patients who died during care was understandably never captured. Finally, due to the wide range of reasons for attending the facility in our sample it was not possible to explore the effect of objective indicators such as adherence to clinical guidelines on perceptions of care despite collecting data on treatment delivery (e.g. tests delivered, observations delivered, consultation etc. – see Supplementary File 2).

In summary, our study has characterised how service users in Malawi perceive the quality of healthcare services they receive across the country. Whilst, generally, quality was perceived as being high this may be attributable to low expectations of care quality driven by the health system’s historic delivery of low-quality services, or unavailability of services, attributable to constrained resources. In addition to addressing gaps in service delivery, such as improving the availability of required medicines, empirical research exploring the drivers of improved perceptions of care quality in CHAM facilities, compared to government facilities, could be leveraged to improve service user-reported quality nationally.

## Supporting information

Supplementary File 2 - Survey questions

Supplementary File 3 - Data cleaning Stata .do file

Supplementary File 4 - Data analysis Stata .do file

## Data Availability

All data produced in the present study are available upon reasonable request to the authors

