## Supplementary File 1 for "Healthcare service user-reported quality of care in Malawi: a national multi-facility cross-sectional study"

**Table S1 - STROBE Statement for cross-sectional studies**

|  | **Item No** | **Recommendation** | **Page No** |
| --- | --- | --- | --- |
| **Title and abstract** | 1 | (*a*) Indicate the study’s design with a commonly used term in the title or the abstract | 1 |
|  |  | (*b*) Provide in the abstract an informative and balanced summary of what was done and what was found | 1-2 |
| **Introduction** | | | |
| Background/rationale | 2 | Explain the scientific background and rationale for the investigation being reported | 3-4 |
| Objectives | 3 | State specific objectives, including any prespecified hypotheses | 4 |
| **Methods** | | | |
| Study design | 4 | Present key elements of study design early in the paper | 4 |
| Setting | 5 | Describe the setting, locations, and relevant dates, including periods of recruitment, exposure, follow-up, and data collection | 4-5 |
| Participants | 6 | (*a*) Give the eligibility criteria, and the sources and methods of selection of participants | 5 |
| Variables | 7 | Clearly define all outcomes, exposures, predictors, potential confounders, and effect modifiers. Give diagnostic criteria, if applicable | 6-7, Table 1 |
| Data sources/ measurement | 8* | For each variable of interest, give sources of data and details of methods of assessment (measurement). Describe comparability of assessment methods if there is more than one group | 5-6 |
| Bias | 9 | Describe any efforts to address potential sources of bias |  |
| Study size | 10 | Explain how the study size was arrived at | 5 |
| Quantitative variables | 11 | Explain how quantitative variables were handled in the analyses. If applicable, describe which groupings were chosen and why | Table 1, Table S2 |
| Statistical methods | 12 | (*a*) Describe all statistical methods, including those used to control for confounding | 6-7 |
|  |  | (*b*) Describe any methods used to examine subgroups and interactions | 6-7 |
|  |  | (*c*) Explain how missing data were addressed | 6 |
|  |  | (*d*) If applicable, describe analytical methods taking account of sampling strategy | 7 |
|  |  | (*e*) Describe any sensitivity analyses | N/A |
| **Results** | | | |
| Participants | 13* | (a) Report numbers of individuals at each stage of study—eg numbers potentially eligible, examined for eligibility, confirmed eligible, included in the study, completing follow-up, and analysed | 7 |
|  |  | (b) Give reasons for non-participation at each stage | N/A |
|  |  | (c) Consider use of a flow diagram | N/A |
| Descriptive data | 14* | (a) Give characteristics of study participants (eg demographic, clinical, social) and information on exposures and potential confounders | 7, Table 2 |
|  |  | (b) Indicate number of participants with missing data for each variable of interest | Table 2 |
| Outcome data | 15* | Report numbers of outcome events or summary measures | 8, Table 3 |
| Main results | 16 | (*a*) Give unadjusted estimates and, if applicable, confounder-adjusted estimates and their precision (eg, 95% confidence interval). Make clear which confounders were adjusted for and why they were included | 8-10, Tables 4 & 5 |
|  |  | (*b*) Report category boundaries when continuous variables were categorized | N/A |
|  |  | (*c*) If relevant, consider translating estimates of relative risk into absolute risk for a meaningful time period | N/A |
| Other analyses | 17 | Report other analyses done—eg analyses of subgroups and interactions, and sensitivity analyses | Tables S4-S7 |
| **Discussion** | | | |
| Key results | 18 | Summarise key results with reference to study objectives | 10 |
| Limitations | 19 | Discuss limitations of the study, taking into account sources of potential bias or imprecision. Discuss both direction and magnitude of any potential bias | 11-12 |
| Interpretation | 20 | Give a cautious overall interpretation of results considering objectives, limitations, multiplicity of analyses, results from similar studies, and other relevant evidence | 12 |
| Generalisability | 21 | Discuss the generalisability (external validity) of the study results | 12 |
| **Other information** | | | |
| Funding | 22 | Give the source of funding and the role of the funders for the present study and, if applicable, for the original study on which the present article is based | / |

*Give information separately for exposed and unexposed groups.

**Note:** An Explanation and Elaboration article discusses each checklist item and gives methodological background and published examples of transparent reporting. The STROBE checklist is best used in conjunction with this article (freely available on the Web sites of PLoS Medicine at http://www.plosmedicine.org/, Annals of Internal Medicine at http://www.annals.org/, and Epidemiology at http://www.epidem.com/). Information on the STROBE Initiative is available at www.strobe-statement.org.

**Table S2 – Included facilities by stratification Type**

| **Facility Name and District** | **Facility Type** | **Facility Ownership** | **Facility Location** | **Patient Volume** |
| --- | --- | --- | --- | --- |
| Lilongwe Central Hospital, Lilongwe | Tertiary Hospital | Government | Urban | High |
| Mzuzu Cental Hospital, Mzuzu | Tertiary Hospital | Government | Urban | High |
| Queen Elizabeth Central Hospital, Blantyre | Tertiary Hospital | Government | Urban | High |
| Zomba Central Hospital, Zomba | Tertiary Hospital | Government | Urban | High |
| Zomba Mental Hospital, Zomba | Tertiary Hospital | Government | Urban | High |
| Bwaila Hospital, Lilongwe | District Hospital | Government | Urban | High |
| Chiradzulu District Hospital, Chiradzulu | District Hospital | Government | Rural | High |
| Nkhata Bay District Hospital, Nkhata Bay | District Hospital | Government | Urban | High |
| Bolero Rural Hospital, Rumphi | Community Hospital | Government | Urban | High |
| Chintheche Rural Hospital, Nkhata Bay | Community Hospital | Government | Urban | Low |
| Daeyang Luke Hospital, Lilongwe | Community Hospital | CHAM | Urban | High |
| Kaporo Rural Hospital, Karonga | Community Hospital | Government | Rural | High |
| Likoma/St Peters, Likoma | Community Hospital | CHAM | Urban | Low |
| Mhuju Rural Hospital, Rumphi | Community Hospital | Government | Rural | Low |
| Mlambe Hospital, Blantyre | Community Hospital | CHAM | Rural | High |
| Mua Mission Hospital, Dedza | Community Hospital | CHAM | Urban | High |
| St Johns Mission Hospital, Blantyre | Community Hospital | CHAM | Urban | Low |
| Trinity - Fatima Hospital, Nsanje | Community Hospital | CHAM | Rural | Low |
| Bilira Health Centre, Ntcheu | Health Centre | Government | Rural | High |
| Chiwe Health Centre, Lilongwe | Health Centre | CHAM | Rural | Low |
| Dzenza Health Centre, Lilongwe | Health Centre | CHAM | Urban | Low |
| Lisungwi Health Centre, Neno | Health Centre | Government | Rural | Low |
| Makwasa Estate Clinic, Thyolo | Health Centre | Private for profit | Urban | Low |
| Malabada Health Centre, Blantyre | Health Centre | CHAM | Urban | High |
| Mianga Estate Clinic, Thyolo | Health Centre | Private for profit | Urban | Low |
| Mlolo Dispensary, Nsanje | Health Centre | Government | Rural | Low |
| Mzuzu Urban Health Centre, Mzimba | Health Centre | Government | Urban | High |
| Pirimiti Health Centre, Zomba | Health Centre | CHAM | Rural | High |
| Sharpe Valley Health Centre, Ntcheu | Health Centre | CHAM | Rural | High |
| Zingwangwa Urban Health Centre, Blantyre | Health Centre | Government | Urban | Low |

**Table S3 – Derivation of select covariates**

| **Covariate** | **Description** | **Survey statement(s) used to derive covariate** | **Derivation method and rationale** |
| --- | --- | --- | --- |
| Age | Service users’ age in ten-year units | Age (years) | Continuous variable calculated as age (years) / 10. Age was rescaled to decade units for clearer interpretation of effect sizes. |
| Education level | Respondent’s education level (none/some primary education, primary/ some secondary education, secondary and above) | Highest education level of respondent (none, some primary, primary completed, some secondary, secondary completed, tertiary) | Categorical variable created by grouping initial categories used in survey statement. None and some primary combined. Primary and some secondary combined. Secondary and tertiary combined.  Catagories combined to reduce model degrees of freedom. |
| Wealth quintile | Respondent’s wealth quintile (1-5) | Do you have the following household assets?  Electricity (yes, no)  Radio (yes, no)  Television (yes, no)  Mobile phone (yes, no)  Non-mobile telephone (yes, no)  Computer (yes, no)  Refrigerator (yes, no)  Koloboyi [cool box] (yes, no)  Paraffin lamp (yes, no)  Torch (yes, no)  Bed with mattress (yes, no)  Sofa set (yes, no)  Ownership of agricultural land (yes, no)  Ownership of farm animals (yes, no) Bicycle (yes, no)  Motorcycle/scooter (yes, no)  Car/truck (yes, no) | Categorical variable created using Principal Component Analysis (PCA) with all assets listed here.  PCA approach allows for generation of single index accounting for assets which is comparable across the population and with other estimates such as the Demographic and Health Survey data. A 5 point categorical variable also allows for a more parsimonious model than including all assets. |
| Illness severity | Self-reported severity of illness for service user (healthy, mild symptoms, moderate symptoms, severe/life-threatening symptoms) | How can you rate the severity of your illness? (healthy, mild symptoms, moderate symptoms, severe symptoms, life-threatening condition) | Categorical variable created by grouping final two categories used in survey statement – Severe symptoms and life-threatening condition  Combined due to small numbers in final two categories. |
| Reason for attendance | Respondent’s reason for attending facility (adult seeking care, adult accompanying a child, adult accompanying an adult) | Age (years)  Reason for attendance? (unwell, accompanying some-one who is unwell, for a test, to receive test results, for medicines, other) | Categorical variable created by using following conditions:  Adults seeking care (reason for attendance (unwell, for a test, to receive test results, for medicines, other) and age > 17)  Adults accompanying an adult (reason for attendance (accompanying some-one who is unwell) and age > 17)  Adults accompanying a child (reason for attendance (accompanying some-one who is unwell) and age < 17)  Variable created to explore variation in perceived quality of care by reason for attendance and age. |
| Facility ownership | Ownership of visited facility (governmental, non-governmental) | Facility ownership (government, CHAM, private for profit) | Binary variable created for non-governmental facilities (CHAM or Private-for-profit (PFP)) and governmental.  CHAM and Private-for-profit facilities combined due to small number of PFP. |
| Travel time to facility | Time taken to reach visited facility for care in ten-minute units | How many minutes does it take you to get to this health facility? | Calculated as time taken to reach facility (minutes) / 10. Travel time was rescaled to ten-minute units for clearer interpretation of effect sizes. |
| Service area | Service area attended during visit to facility (disease clinics, emergency department, maternity or gynaecology inpatients, maternity or gynaecology outpatients, other outpatients, other inpatients, surgery) | Clinic (Outpatients – children, Outpatients – adults, Outpatients – general, HIV clinic, TB clinic, Malaria clinic, Cervical cancer, Antenatal care clinic, Antenatal care ward, OBGYN clinic, OBGYN ward, Intrapartum and immediate newborn care, Postnatal ward ,Postnatal care contact (maternal), Postnatal care contact (neonatal), Post abortion care, Ectopic Pregnancy case management, NCD clinics/inpatient, Intensive and emergency care and recovery room (adult/paediatric), Surgical Clinic, Surgical Ward, Hypertension clinic, Epilepsy clinic, Maternity (labour and delivery) ward, Neonatal ward, Paediatric (children) ward, Male ward, Female ward, Neonatal intensive care (NICU), Paediatric intensive care (PICU), High Dependency / Intensive Care Unit, General emergency unit, Paediatric emergency unit, Diabetes clinic, Surgery (operating theatre), Dental Clinic, Eye Clinic, Pharmacy, Laboratory, Radiology, Dermatology, STI Clinic, Oncology Clinic, Physiotherapy Clinic) | Categorical variable derived by grouping initial categories as follows:  Disease clinics (NCD clinics, Cancer clinic, Dermatology, Diabetes clinic, Epilepsy clinic, Hypertension clinic, Oncology Clinic, Dental Clinic, Eye Clinic, HIV clinic, TB clinic, STI Clinic, NCD clinics/inpatient)  Other clinics/outpatients (Outpatients - adults, Outpatients - children, Outpatients – general, Physiotherapy Clinic, Radiology, Laboratory)  Maternity / Gynaecology outpatient, (Antenatal care clinic, Ectopic Pregnancy case management, Post abortion care, Postnatal care contact (maternal), OBGYN clinic, Cervical cancer, Postnatal care contact (neonatal))  Maternity / Gynaecology inpatient, (Antenatal care ward, Maternity (labour and delivery) ward, OBGYN ward, Postnatal ward)  Other inpatient ward, (Male ward, Female ward, Neonatal ward, Paediatric (children) ward, High Dependency / Intensive Care Unit, Intensive and emergency care and recovery room (adult/paediatric), Neonatal intensive care (NICU), Paediatric intensive care (PICU))  Emergency department (General emergency unit, Paediatric emergency unit)  Surgery (Surgery (all), Surgery (operating theatre), Surgical Ward, Surgical Clinic)  These groupings were developed to due to small numbers of participants for many clinics and to reduce to reduce model degrees of freedom. |
| Time spent with healthcare worker (HCW) | Self-reported total time spent with any healthcare workers during visit to facility in ten-minute units | Health worker 1…(i)th - Who did you see today?  How many minutes did you spend with the health worker? | Continuous variable derived by summing the total reported time respondents believed they had spent with each healthcare worker they recorded /10. Time was rescaled to ten-minute units for clearer interpretation of effect sizes. |
| Any fees paid to the facility | Whether any fees were paid directly to the health facility during this visit (yes, no) | Any payment made to facility (MWK)? | Binary variable created from continuous variable in which a value of 0 MWK is categorised as 0 and a value above 0 MWK is categorised as 1.  Binary variable created to aid interpretation when used as covariate. |
| Any inpatient days | Whether the service user spent any days at the facility as an inpatient (yes, no) | For how many days have you stayed at the facility? | Binary variable created from continuous variable in which a value of 0 days is categorised as 0 and a value above 0 days is categorised as 1  Binary variable created to aid interpretation when used as covariate |

**Table S4 – Multivariable analysis of individual and health service factors and service user reported quality of care relating to healthcare worker consultation**

|  | **Service user reported Quality of Care - Healthcare Worker Consultation**  Adjusted Relative Risk Ratio (ARRR) (95% CI)  (Reference outcome - Good) | | | | | |
| --- | --- | --- | --- | --- | --- | --- |
|  | **Model 1** | | **Model 2** | | **Model 3** | |
|  | **Patient and illness characteristics** | | **Model 1 + Facility and visit characteristics** | | **Model 2 + 3** | |
| VARIABLES | Neutral - Very bad | Very Good | Neutral - Very bad | Very Good | Neutral - Very bad | Very Good |
| **Age** (10-year units) | 1.008 | 1.084 | 0.988 | 1.060 | 0.978 | 1.058 |
|  | (0.885 - 1.149) | (0.955 - 1.231) | (0.876 - 1.113) | (0.937 - 1.199) | (0.855 - 1.118) | (0.946 - 1.183) |
| **Sex** (male) | 0.933 | 1.189 | 1.003 | 1.298*** | 0.972 | 1.287** |
|  | (0.626 - 1.391) | (0.969 - 1.459) | (0.657 - 1.533) | (1.082 - 1.556) | (0.597 - 1.581) | (1.057 - 1.567) |
| **Education level** *(Ref – No education/some primary)*  *Primary/some secondary* | 0.597 | 0.991 | 0.642 | 0.958 | 0.703 | 0.940 |
|  | (0.308 - 1.159) | (0.697 - 1.409) | (0.356 - 1.158) | (0.688 - 1.336) | (0.360 - 1.374) | (0.679 - 1.301) |
| *Secondary/tertiary* | 0.648 | 1.627 | 0.716 | 1.320 | 0.837 | 1.426 |
|  | (0.333 - 1.263) | (0.959 - 2.761) | (0.395 - 1.297) | (0.808 - 2.156) | (0.449 - 1.560) | (0.880 - 2.312) |
| **Wealth quintile** *(Ref – First quintile)*  *Second quintile* | 1.176 | 1.186 | 1.106 | 1.084 | 2.152 | 0.964 |
|  | (0.481 - 2.876) | (0.827 - 1.700) | (0.427 - 2.865) | (0.757 - 1.553) | (0.928 - 4.992) | (0.693 - 1.342) |
| *Third quintile* | 1.186 | 1.074 | 1.298 | 0.980 | 2.000 | 0.871 |
|  | (0.528 - 2.666) | (0.711 - 1.624) | (0.580 - 2.905) | (0.666 - 1.443) | (0.703 - 5.689) | (0.614 - 1.236) |
| *Fourth quintile* | 1.596 | 1.243 | 1.512 | 0.974 | 3.158*** | 0.927 |
|  | (0.713 - 3.574) | (0.736 - 2.101) | (0.678 - 3.371) | (0.594 - 1.596) | (1.424 - 7.003) | (0.609 - 1.410) |
| *Fifth quintile* | 1.770 | 1.413 | 1.542 | 1.037 | 3.345** | 0.951 |
|  | (0.649 - 4.829) | (0.800 - 2.498) | (0.514 - 4.625) | (0.633 - 1.697) | (1.314 - 8.516) | (0.603 - 1.499) |
| **Illness severity** *(Ref – Healthy)*  *Mild symptoms* | 2.036 | 0.229*** | 2.637 | 0.208*** | 1.359 | 0.231*** |
|  | (0.484 - 8.569) | (0.113 - 0.464) | (0.601 - 11.57) | (0.103 - 0.421) | (0.412 - 4.483) | (0.102 - 0.522) |
| *Moderate symptoms* | 2.393 | 0.193*** | 3.124 | 0.187*** | 2.036 | 0.202*** |
|  | (0.647 - 8.848) | (0.103 - 0.365) | (0.742 - 13.15) | (0.0852 - 0.409) | (0.518 - 8.008) | (0.0797 - 0.510) |
| *Severe/life threatening symptoms* | 6.207** | 0.291*** | 7.547*** | 0.320** | 4.061 | 0.348 |
|  | (1.469 - 26.23) | (0.125 - 0.675) | (1.643 - 34.67) | (0.121 - 0.845) | (0.945 - 17.44) | (0.111 - 1.091) |
| **Previously sought care** (yes) | 1.686 | 0.505*** | 1.749 | 0.495*** | 2.097** | 0.451*** |
|  | (0.892 - 3.187) | (0.341 - 0.746) | (0.849 - 3.603) | (0.321 - 0.766) | (1.089 - 4.039) | (0.285 - 0.714) |
| **Referral** (yes) | 1.527 | 3.286** | 1.894 | 1.755** | 1.792 | 1.702*** |
|  | (0.584 - 3.995) | (1.030 - 10.49) | (0.664 - 5.401) | (1.091 - 2.824) | (0.637 - 5.041) | (1.175 - 2.464) |
| **Facility type** *(Ref – Health centre)*  *Community hospital* |  |  | 0.286*** | 1.817 | 0.313*** | 1.233 |
|  |  |  | (0.114 - 0.720) | (0.617 - 5.351) | (0.159 - 0.616) | (0.399 - 3.814) |
| *District Hospital* |  |  | 0.0574*** | 0.918 | 0.100*** | 0.886 |
|  |  |  | (0.0141 - 0.234) | (0.126 - 6.690) | (0.0316 - 0.319) | (0.133 - 5.878) |
| *Tertiary hospital* |  |  | 0.254** | 5.778** | 0.385** | 5.596 |
|  |  |  | (0.0817 - 0.792) | (1.045 - 31.96) | (0.183 - 0.808) | (0.849 - 36.87) |
| **Facility ownership** (non-governmental) |  |  | 0.498 | 2.656 | 1.376 | 5.373** |
|  |  |  | (0.160 - 1.554) | (0.719 - 9.807) | (0.543 - 3.485) | (1.354 - 21.33) |
| **Facility location** (rural) |  |  | 0.670 | 0.880 | 0.619 | 0.810 |
|  |  |  | (0.233 - 1.924) | (0.356 - 2.172) | (0.281 - 1.364) | (0.285 - 2.306) |
| **Service area** *(Ref – Other clinics / outpatients))*  *Disease clinics* |  |  | 1.598 | 1.084 | 1.684 | 1.049 |
|  |  |  | (0.689 - 3.704) | (0.588 - 2.000) | (0.703 - 4.032) | (0.598 - 1.840) |
| *Emergency department* |  |  | 3.753** | 0.886 | 3.494** | 0.766 |
|  |  |  | (1.204 - 11.70) | (0.228 - 3.439) | (1.172 - 10.42) | (0.190 - 3.093) |
| *Maternity/Gynae inpatient* |  |  | 7.213*** | 3.065 | 6.300** | 1.970 |
|  |  |  | (1.898 - 27.42) | (0.823 - 11.42) | (1.479 - 26.84) | (0.433 - 8.965) |
| *Maternity/Gynae outpatient* |  |  | 0.256 | 1.195 | 0.363 | 1.205 |
|  |  |  | (0.0287 - 2.284) | (0.609 - 2.343) | (0.0519 - 2.533) | (0.559 - 2.596) |
| *Other inpatient ward* |  |  | 0.553 | 0.436 | 0.472 | 0.239** |
|  |  |  | (0.197 - 1.550) | (0.121 - 1.572) | (0.146 - 1.529) | (0.0647 - 0.879) |
| *Surgery* |  |  | 2.415 | 4.716*** | 1.290 | 3.688** |
|  |  |  | (0.277 - 21.09) | (1.481 - 15.02) | (0.0992 - 16.79) | (1.292 - 10.53) |
| **Any inpatient days** (yes) |  |  | 0.943 | 2.238 | 1.728 | 2.469 |
|  |  |  | (0.345 - 2.578) | (0.716 - 6.990) | (0.602 - 4.964) | (0.805 - 7.574) |
| **Travel time to facility** (10-minute units) |  |  |  |  | 1.049*** | 1.023 |
|  |  |  |  |  | (1.012 - 1.088) | (0.996 - 1.051) |
| **Time spent with HCW** (10-minute units) |  |  |  |  | 0.943 | 1.069 |
|  |  |  |  |  | (0.757 - 1.173) | (0.979 - 1.168) |
| **Accessed required medication** *(Ref – Yes)*  *No* |  |  |  |  | 3.755*** | 0.573 |
|  |  |  |  |  | (1.974 - 7.142) | (0.263 - 1.248) |
| *None prescribed* |  |  |  |  | 0.390 | 1.211 |
|  |  |  |  |  | (0.0507 - 3.005) | (0.682 - 2.150) |
| **Any fees paid to the facility** (Yes) |  |  |  |  | 0.284** | 0.609 |
|  |  |  |  |  | (0.0982 - 0.822) | (0.264 - 1.405) |
| Constant | 0.0136*** | 1.506 | 0.0336*** | 0.597 | 0.00448*** | 0.674 |
|  | (0.00338 - 0.0549) | (0.554 - 4.096) | (0.00694 - 0.162) | (0.112 - 3.193) | (0.000579 - 0.0346) | (0.0844 - 5.384) |
| Observations | 4,086 | 4,086 | 4,086 | 4,086 | 3,613 | 3,613 |
| CI in parentheses. *** p<0.01, ** p<0.05, * p<0.1 | | | | | | |

**Table S5 – Multivariable analysis of individual and health service factors and service user reported quality of care relating to treatment with respect**

|  | **Service user reported Quality of Care – Treated with Respect**  Adjusted Relative Risk Ratio (ARRR) (95% CI)  (Reference outcome - Good) | | | | | |
| --- | --- | --- | --- | --- | --- | --- |
|  | **Model 1** | | **Model 2** | | **Model 3** | |
|  | **Patient and illness characteristics** | | **Model 1 + Facility and visit characteristics** | | **Model 2 + Additional service delivery factors** | |
| VARIABLES | Neutral - Very bad | Very Good | Neutral - Very bad | Very Good | Neutral - Very bad | Very Good |
| **Age** (10-year units) | 1.043 | 1.091 | 0.968 | 1.064 | 0.966 | 1.063 |
|  | (0.888 - 1.226) | (0.964 - 1.233) | (0.830 - 1.129) | (0.944 - 1.199) | (0.835 - 1.119) | (0.951 - 1.188) |
| **Sex** (male) | 0.937 | 1.169 | 0.942 | 1.281*** | 1.017 | 1.312*** |
|  | (0.654 - 1.343) | (0.963 - 1.419) | (0.666 - 1.333) | (1.079 - 1.521) | (0.720 - 1.437) | (1.092 - 1.576) |
| **Education level** *(Ref – No education/some primary)*  *Primary/some secondary* | 0.971 | 0.915 | 0.981 | 0.906 | 1.040 | 0.916 |
|  | (0.624 - 1.511) | (0.649 - 1.290) | (0.618 - 1.556) | (0.654 - 1.253) | (0.666 - 1.625) | (0.670 - 1.252) |
| *Secondary/tertiary* | 0.819 | 1.490 | 0.826 | 1.257 | 0.863 | 1.425 |
|  | (0.491 - 1.365) | (0.880 - 2.523) | (0.482 - 1.415) | (0.784 - 2.015) | (0.468 - 1.591) | (0.893 - 2.274) |
| **Wealth quintile** *(Ref – First quintile)*  *Second quintile* | 1.325 | 1.136 | 1.275 | 1.034 | 1.534 | 0.942 |
|  | (0.649 - 2.706) | (0.816 - 1.582) | (0.602 - 2.701) | (0.740 - 1.446) | (0.613 - 3.843) | (0.683 - 1.298) |
| *Third quintile* | 1.564 | 1.024 | 1.447 | 0.963 | 1.677 | 0.885 |
|  | (0.831 - 2.945) | (0.690 - 1.522) | (0.776 - 2.696) | (0.665 - 1.396) | (0.768 - 3.660) | (0.629 - 1.246) |
| *Fourth quintile* | 1.422 | 1.129 | 1.296 | 0.924 | 1.495 | 0.853 |
|  | (0.697 - 2.900) | (0.677 - 1.882) | (0.671 - 2.505) | (0.558 - 1.529) | (0.632 - 3.534) | (0.544 - 1.337) |
| *Fifth quintile* | 2.048** | 1.296 | 1.686* | 0.980 | 1.893 | 0.901 |
|  | (1.035 - 4.055) | (0.751 - 2.236) | (0.938 - 3.029) | (0.597 - 1.609) | (0.886 - 4.042) | (0.557 - 1.456) |
| **Illness severity** *(Ref – Healthy)*  *Mild symptoms* | 0.391*** | 0.261*** | 0.442 | 0.292*** | 0.514 | 0.300*** |
|  | (0.212 - 0.719) | (0.133 - 0.514) | (0.184 - 1.063) | (0.144 - 0.593) | (0.178 - 1.485) | (0.132 - 0.680) |
| *Moderate symptoms* | 0.489** | 0.207*** | 0.661 | 0.245*** | 0.743 | 0.248*** |
|  | (0.242 - 0.987) | (0.111 - 0.386) | (0.249 - 1.757) | (0.118 - 0.509) | (0.252 - 2.193) | (0.0993 - 0.618) |
| *Severe/life threatening symptoms* | 0.913 | 0.324*** | 1.325 | 0.427 | 0.975 | 0.457 |
|  | (0.359 - 2.322) | (0.146 - 0.719) | (0.443 - 3.963) | (0.177 - 1.027) | (0.262 - 3.630) | (0.152 - 1.371) |
| **Previously sought care** (yes) | 1.468 | 0.529*** | 1.236 | 0.511*** | 1.175 | 0.479*** |
|  | (0.849 - 2.538) | (0.369 - 0.757) | (0.781 - 1.955) | (0.342 - 0.765) | (0.753 - 1.835) | (0.309 - 0.743) |
| **Referral** (yes) | 1.714 | 3.108 | 1.418 | 1.864** | 1.131 | 1.712** |
|  | (0.853 - 3.442) | (0.928 - 10.42) | (0.649 - 3.099) | (1.105 - 3.144) | (0.599 - 2.138) | (1.089 - 2.692) |
| **Facility type** *(Ref – Health centre)*  *Community hospital* |  |  | 0.441*** | 1.266 | 0.423** | 1.044 |
|  |  |  | (0.239 - 0.813) | (0.429 - 3.736) | (0.213 - 0.840) | (0.361 - 3.016) |
| *District Hospital* |  |  | 0.523 | 0.512 | 0.568 | 0.677 |
|  |  |  | (0.160 - 1.703) | (0.0623 - 4.199) | (0.183 - 1.760) | (0.110 - 4.149) |
| *Tertiary hospital* |  |  | 0.762 | 3.273 | 1.169 | 4.190 |
|  |  |  | (0.261 - 2.226) | (0.536 - 19.99) | (0.484 - 2.825) | (0.678 - 25.89) |
| **Facility ownership** (non-governmental) |  |  | 0.731 | 1.914 | 1.467 | 4.607** |
|  |  |  | (0.406 - 1.315) | (0.467 - 7.840) | (0.656 - 3.281) | (1.162 - 18.26) |
| **Facility location** (rural) |  |  | 0.576 | 0.945 | 0.682 | 0.760 |
|  |  |  | (0.267 - 1.243) | (0.378 - 2.360) | (0.286 - 1.630) | (0.278 - 2.077) |
| **Service area** *(Ref – Other clinics / outpatients))*  *Disease clinics* |  |  | 2.276** | 1.250 | 2.207 | 1.230 |
|  |  |  | (1.039 - 4.988) | (0.701 - 2.230) | (0.974 - 5.002) | (0.714 - 2.118) |
| *Emergency department* |  |  | 4.887** | 0.955 | 4.034** | 0.777 |
|  |  |  | (1.368 - 17.46) | (0.299 - 3.053) | (1.186 - 13.72) | (0.236 - 2.559) |
| *Maternity/Gynae inpatient* |  |  | 4.809** | 3.894** | 2.113 | 2.459 |
|  |  |  | (1.197 - 19.31) | (1.033 - 14.68) | (0.513 - 8.696) | (0.563 - 10.73) |
| *Maternity/Gynae outpatient* |  |  | 1.266 | 1.342 | 0.963 | 1.421 |
|  |  |  | (0.608 - 2.638) | (0.671 - 2.681) | (0.457 - 2.033) | (0.660 - 3.059) |
| *Other inpatient ward* |  |  | 1.222 | 0.513 | 0.751 | 0.261** |
|  |  |  | (0.591 - 2.525) | (0.138 - 1.910) | (0.301 - 1.874) | (0.0722 - 0.946) |
| *Surgery* |  |  | 5.420** | 5.264*** | 3.129 | 4.126*** |
|  |  |  | (1.493 - 19.67) | (1.789 - 15.48) | (0.756 - 12.96) | (1.621 - 10.50) |
| **Any inpatient days** (yes) |  |  | 0.587 | 1.684 | 0.915 | 2.077 |
|  |  |  | (0.255 - 1.348) | (0.492 - 5.763) | (0.450 - 1.862) | (0.672 - 6.420) |
| **Travel time to facility** (10-minute units) |  |  |  |  | 1.028 | 1.021 |
|  |  |  |  |  | (0.991 - 1.066) | (0.991 - 1.051) |
| **Time spent with HCW** (10-minute uints) |  |  |  |  | 1.056 | 1.081 |
|  |  |  |  |  | (0.950 - 1.173) | (0.989 - 1.181) |
| **Accessed required medication** *(Ref – Yes)*  *No* |  |  |  |  | 3.421*** | 0.557 |
|  |  |  |  |  | (1.977 - 5.920) | (0.298 - 1.039) |
| *None prescribed* |  |  |  |  | 2.479 | 1.033 |
|  |  |  |  |  | (0.883 - 6.959) | (0.592 - 1.802) |
| **Any fees paid to the facility** (Yes) |  |  |  |  | 0.396** | 0.602 |
|  |  |  |  |  | (0.190 - 0.824) | (0.269 - 1.349) |
| Constant | 0.0460*** | 1.655 | 0.0880*** | 0.864 | 0.0851*** | 0.637 |
|  | (0.0180 - 0.118) | (0.641 - 4.274) | (0.0307 - 0.252) | (0.172 - 4.336) | (0.0221 - 0.327) | (0.0872 - 4.658) |
| Observations | 4,088 | 4,088 | 4,088 | 4,088 | 3,620 | 3,620 |
| CI in parentheses. *** p<0.01, ** p<0.05 | | | | | | |

**Table S6 – Multivariable analysis of individual and health service factors and service user reported quality of care relating to waiting times**

|  | **Service user reported Quality of Care – Waiting Times**  Adjusted Relative Risk Ratio (ARRR) (95% CI)  (Reference outcome - Good) | | | | | |
| --- | --- | --- | --- | --- | --- | --- |
|  | **Model 1** | | **Model 2** | | **Model 3** | |
|  | **Patient and illness characteristics** | | **Model 1 + Facility and visit characteristics** | | **Model 2 + Additional service delivery factors** | |
| VARIABLES | Neutral - Very bad | Very Good | Neutral - Very bad | Very Good | Neutral - Very bad | Very Good |
| Age (10-year units) | 1.087 | 1.060 | 1.059 | 1.050 | 1.070 | 1.042 |
|  | (0.986 - 1.198) | (0.937 - 1.198) | (0.991 - 1.132) | (0.934 - 1.180) | (0.992 - 1.154) | (0.940 - 1.156) |
| **Sex** (Male) | 1.042 | 1.093 | 1.078 | 1.280*** | 1.210 | 1.271** |
|  | (0.874 - 1.242) | (0.867 - 1.378) | (0.905 - 1.284) | (1.068 - 1.535) | (0.998 - 1.467) | (1.049 - 1.540) |
| **Education level** *(Ref – No education/some primary)*  *Primary/some secondary* | 0.736** | 1.095 | 0.777 | 1.122 | 0.763 | 1.063 |
|  | (0.557 - 0.974) | (0.778 - 1.541) | (0.584 - 1.034) | (0.835 - 1.507) | (0.571 - 1.021) | (0.801 - 1.411) |
| *Secondary/tertiary* | 0.927 | 1.745** | 0.980 | 1.441 | 1.018 | 1.525** |
|  | (0.611 - 1.405) | (1.063 - 2.864) | (0.640 - 1.500) | (0.921 - 2.254) | (0.637 - 1.627) | (1.006 - 2.312) |
| **Wealth quintile** *(Ref – First quintile)*  *Second quintile* | 0.864 | 1.235 | 0.801 | 1.152 | 0.838 | 1.089 |
|  | (0.588 - 1.268) | (0.932 - 1.638) | (0.552 - 1.161) | (0.892 - 1.487) | (0.548 - 1.281) | (0.837 - 1.417) |
| *Third quintile* | 0.704 | 1.016 | 0.707** | 0.927 | 0.814 | 0.857 |
|  | (0.495 - 1.001) | (0.673 - 1.535) | (0.522 - 0.956) | (0.646 - 1.331) | (0.569 - 1.164) | (0.599 - 1.228) |
| *Fourth quintile* | 0.852 | 1.194 | 0.824 | 0.961 | 0.883 | 0.903 |
|  | (0.587 - 1.238) | (0.747 - 1.907) | (0.569 - 1.194) | (0.637 - 1.450) | (0.586 - 1.332) | (0.610 - 1.337) |
| *Fifth quintile* | 0.865 | 1.429 | 0.759 | 1.042 | 0.833 | 0.965 |
|  | (0.545 - 1.371) | (0.807 - 2.531) | (0.520 - 1.109) | (0.644 - 1.684) | (0.605 - 1.147) | (0.589 - 1.580) |
| **Illness severity** *(Ref – Healthy)*  *Mild symptoms* | 0.592* | 0.311*** | 0.712 | 0.382*** | 0.746 | 0.465** |
|  | (0.325 - 1.076) | (0.161 - 0.601) | (0.447 - 1.132) | (0.214 - 0.680) | (0.411 - 1.352) | (0.239 - 0.906) |
| *Moderate symptoms* | 0.562* | 0.274*** | 0.749 | 0.409*** | 0.784 | 0.474** |
|  | (0.291 - 1.085) | (0.154 - 0.490) | (0.435 - 1.289) | (0.234 - 0.715) | (0.408 - 1.508) | (0.246 - 0.914) |
| *Severe/life threatening symptoms* | 0.547 | 0.393** | 0.732 | 0.712 | 0.589 | 0.849 |
|  | (0.247 - 1.212) | (0.176 - 0.878) | (0.389 - 1.379) | (0.332 - 1.525) | (0.261 - 1.330) | (0.364 - 1.979) |
| **Previously sought care** (yes) | 1.204 | 0.562*** | 1.051 | 0.593** | 0.895 | 0.576** |
|  | (0.823 - 1.762) | (0.378 - 0.833) | (0.719 - 1.537) | (0.398 - 0.885) | (0.574 - 1.395) | (0.376 - 0.881) |
| **Referral** (yes) | 1.331 | 2.865 | 1.434 | 1.389 | 1.158 | 1.370** |
|  | (0.435 - 4.079) | (0.909 - 9.025) | (0.833 - 2.468) | (0.941 - 2.050) | (0.695 - 1.930) | (1.009 - 1.860) |
| **Facility type** *(Ref – Health centre)*  *Community hospital* |  |  | 0.509 | 1.393 | 0.553 | 1.021 |
|  |  |  | (0.212 - 1.223) | (0.493 - 3.939) | (0.231 - 1.326) | (0.293 - 3.553) |
| *District Hospital* |  |  | 0.254** | 1.652 | 0.270** | 1.471 |
|  |  |  | (0.0850 - 0.760) | (0.282 - 9.680) | (0.0917 - 0.797) | (0.277 - 7.799) |
| *Tertiary hospital* |  |  | 0.699 | 5.313*** | 1.030 | 4.421** |
|  |  |  | (0.223 - 2.197) | (1.502 - 18.80) | (0.372 - 2.852) | (1.032 - 18.95) |
| **Facility ownership** (non-governmental) |  |  | 0.545 | 5.420*** | 0.941 | 8.298*** |
|  |  |  | (0.256 - 1.159) | (1.772 - 16.57) | (0.355 - 2.497) | (2.425 - 28.39) |
| **Facility location** (rural) |  |  | 0.735 | 0.619 | 0.730 | 0.565 |
|  |  |  | (0.391 - 1.379) | (0.263 - 1.458) | (0.365 - 1.462) | (0.212 - 1.503) |
| **Service area** *(Ref – Other clinics / outpatients))*  *Disease clinics* |  |  | 0.978 | 1.275 | 0.810 | 1.230 |
|  |  |  | (0.646 - 1.482) | (0.822 - 1.979) | (0.536 - 1.223) | (0.805 - 1.879) |
| *Emergency department* |  |  | 1.472 | 1.086 | 1.195 | 0.989 |
|  |  |  | (0.486 - 4.463) | (0.361 - 3.272) | (0.440 - 3.246) | (0.332 - 2.944) |
| *Maternity/Gynae inpatient* |  |  | 2.603 | 2.913** | 1.980 | 2.076 |
|  |  |  | (0.759 - 8.931) | (1.135 - 7.472) | (0.555 - 7.060) | (0.693 - 6.215) |
| *Maternity/Gynae outpatient* |  |  | 1.676 | 1.440 | 1.731 | 1.451 |
|  |  |  | (0.831 - 3.380) | (0.818 - 2.537) | (0.798 - 3.753) | (0.726 - 2.902) |
| *Other inpatient ward* |  |  | 0.383*** | 0.488 | 0.229*** | 0.276** |
|  |  |  | (0.212 - 0.690) | (0.158 - 1.512) | (0.115 - 0.454) | (0.0939 - 0.811) |
| *Surgery* |  |  | 5.580*** | 3.092*** | 3.390*** | 2.576*** |
|  |  |  | (2.866 - 10.86) | (1.609 - 5.940) | (1.854 - 6.197) | (1.339 - 4.955) |
| **Any inpatient days** (yes) |  |  | 0.447** | 3.435** | 0.427** | 3.385*** |
|  |  |  | (0.212 - 0.945) | (1.334 - 8.848) | (0.183 - 0.996) | (1.343 - 8.534) |
| **Travel time to facility** (10-minute units) |  |  |  |  | 1.035** | 1.018 |
|  |  |  |  |  | (1.001 - 1.069) | (0.989 - 1.048) |
| **Time spent with HCW** (10-minute units) |  |  |  |  | 1.103*** | 1.101** |
|  |  |  |  |  | (1.035 - 1.175) | (1.019 - 1.190) |
| **Accessed required medication** *(Ref – Yes)*  *No* |  |  |  |  | 2.153*** | 0.382*** |
|  |  |  |  |  | (1.476 - 3.142) | (0.221 - 0.662) |
| *None prescribed* |  |  |  |  | 0.747 | 1.003 |
|  |  |  |  |  | (0.424 - 1.318) | (0.587 - 1.715) |
| **Any fees paid to the facility** (Yes) |  |  |  |  | 0.327*** | 0.696 |
|  |  |  |  |  | (0.202 - 0.528) | (0.340 - 1.423) |
| Constant | 0.436* | 0.978 | 0.895 | 0.186** | 0.365 | 0.177** |
|  | (0.181 - 1.047) | (0.376 - 2.542) | (0.376 - 2.130) | (0.0467 - 0.745) | (0.109 - 1.224) | (0.0389 - 0.804) |
| Observations | 4,078 | 4,078 | 4,078 | 4,078 | 3,609 | 3,609 |
| CI in parentheses. *** p<0.01, ** p<0.05 | | | | | | |

**Table S7 – Multivariable analysis of individual and health service factors and service user reported quality of care relating to treatment availability**

|  | **Service user reported Quality of Care – Treatment Availability**  Adjusted Relative Risk Ratio (ARRR) (95% CI)  (Reference outcome - Good) | | | | | |
| --- | --- | --- | --- | --- | --- | --- |
|  | **Model 1** | | **Model 2** | | **Model 3** | |
|  | **Patient and illness characteristics** | | **Model 1 + Facility and visit characteristics** | | **Model 2 + Additional service delivery factors** | |
| VARIABLES | Neutral - Very bad | Very Good | Neutral - Very bad | Very Good | Neutral - Very bad | Very Good |
| **Age** (10-year units) | 1.022 | 1.049 | 0.998 | 1.022 | 1.023 | 1.027 |
|  | (0.904 - 1.156) | (0.934 - 1.179) | (0.913 - 1.090) | (0.916 - 1.141) | (0.934 - 1.120) | (0.926 - 1.139) |
| **Sex** (Male) | 1.106 | 1.103 | 1.053 | 1.176 | 1.342** | 1.167 |
|  | (0.865 - 1.414) | (0.910 - 1.335) | (0.822 - 1.348) | (0.982 - 1.407) | (1.041 - 1.730) | (0.964 - 1.413) |
| **Education level** *(Ref – No education/some primary)*  *Primary/some secondary* | 0.803 | 0.959 | 0.848 | 0.968 | 0.762 | 0.974 |
|  | (0.606 - 1.064) | (0.706 - 1.302) | (0.638 - 1.127) | (0.732 - 1.281) | (0.543 - 1.070) | (0.742 - 1.278) |
| *Secondary/tertiary* | 1.023 | 1.527 | 1.167 | 1.294 | 0.991 | 1.413 |
|  | (0.650 - 1.611) | (0.919 - 2.535) | (0.705 - 1.933) | (0.829 - 2.019) | (0.614 - 1.598) | (0.919 - 2.172) |
| **Wealth quintile** *(Ref – First quintile)*  *Second quintile* | 1.000 | 1.203 | 1.010 | 1.117 | 1.150 | 1.063 |
|  | (0.771 - 1.297) | (0.864 - 1.676) | (0.784 - 1.300) | (0.816 - 1.528) | (0.802 - 1.649) | (0.745 - 1.516) |
| *Third quintile* | 0.876 | 1.105 | 0.904 | 1.059 | 0.991 | 0.976 |
|  | (0.683 - 1.123) | (0.729 - 1.677) | (0.698 - 1.170) | (0.724 - 1.548) | (0.760 - 1.291) | (0.682 - 1.398) |
| *Fourth quintile* | 0.917 | 1.176 | 1.014 | 0.984 | 1.577*** | 0.941 |
|  | (0.682 - 1.233) | (0.724 - 1.908) | (0.772 - 1.334) | (0.610 - 1.589) | (1.166 - 2.133) | (0.611 - 1.450) |
| *Fifth quintile* | 0.884 | 1.393 | 0.889 | 1.057 | 1.059 | 1.020 |
|  | (0.615 - 1.270) | (0.782 - 2.483) | (0.628 - 1.257) | (0.638 - 1.753) | (0.772 - 1.453) | (0.609 - 1.708) |
| **Illness severity** *(Ref – Healthy)*  *Mild symptoms* | 1.308 | 0.263*** | 1.264 | 0.284*** | 0.936 | 0.295*** |
|  | (0.623 - 2.746) | (0.142 - 0.487) | (0.587 - 2.722) | (0.150 - 0.540) | (0.410 - 2.136) | (0.139 - 0.623) |
| *Moderate symptoms* | 1.325 | 0.199*** | 1.474 | 0.241*** | 1.686 | 0.255*** |
|  | (0.627 - 2.800) | (0.111 - 0.356) | (0.677 - 3.206) | (0.123 - 0.472) | (0.689 - 4.127) | (0.111 - 0.585) |
| *Severe/life threatening symptoms* | 2.610** | 0.308*** | 3.379*** | 0.424** | 3.144** | 0.423 |
|  | (1.168 - 5.830) | (0.144 - 0.658) | (1.387 - 8.232) | (0.186 - 0.969) | (1.133 - 8.723) | (0.148 - 1.213) |
| **Previously sought care** (yes) | 1.580 | 0.598*** | 1.309 | 0.572*** | 1.354 | 0.534*** |
|  | (0.996 - 2.505) | (0.430 - 0.831) | (0.903 - 1.896) | (0.391 - 0.835) | (0.938 - 1.954) | (0.359 - 0.796) |
| **Referral** (yes) | 1.737 | 2.947 | 2.294 | 1.644 | 1.643 | 1.494** |
|  | (0.576 - 5.237) | (0.885 - 9.810) | (0.985 - 5.342) | (0.980 - 2.759) | (0.707 - 3.816) | (1.001 - 2.229) |
| **Facility type** *(Ref – Health centre)*  *Community hospital* |  |  | 0.649 | 1.185 | 0.749 | 0.955 |
|  |  |  | (0.372 - 1.130) | (0.405 - 3.466) | (0.494 - 1.134) | (0.313 - 2.909) |
| *District Hospital* |  |  | 0.481*** | 0.550 | 0.531 | 0.700 |
|  |  |  | (0.278 - 0.832) | (0.0671 - 4.515) | (0.262 - 1.073) | (0.109 - 4.495) |
| *Tertiary hospital* |  |  | 0.501 | 3.154 | 1.251 | 3.925 |
|  |  |  | (0.179 - 1.403) | (0.571 - 17.43) | (0.638 - 2.451) | (0.666 - 23.15) |
| **Facility ownership** (non-governmental) |  |  | 0.652 | 2.400 | 0.973 | 5.935*** |
|  |  |  | (0.334 - 1.273) | (0.644 - 8.949) | (0.563 - 1.680) | (1.603 - 21.97) |
| **Facility location** (rural) |  |  | 0.582** | 0.825 | 0.931 | 0.710 |
|  |  |  | (0.353 - 0.960) | (0.341 - 1.994) | (0.597 - 1.453) | (0.257 - 1.958) |
| **Service area** *(Ref – Other clinics / outpatients))*  *Disease clinics* |  |  | 1.288 | 1.516 | 1.082 | 1.411 |
|  |  |  | (0.861 - 1.925) | (0.916 - 2.507) | (0.764 - 1.531) | (0.892 - 2.232) |
| *Emergency department* |  |  | 1.033 | 1.211 | 0.723 | 0.986 |
|  |  |  | (0.431 - 2.479) | (0.333 - 4.410) | (0.282 - 1.852) | (0.259 - 3.757) |
| *Maternity/Gynae inpatient* |  |  | 1.353 | 3.336* | 0.927 | 2.499 |
|  |  |  | (0.416 - 4.400) | (0.940 - 11.84) | (0.248 - 3.464) | (0.605 - 10.32) |
| *Maternity/Gynae outpatient* |  |  | 0.763 | 1.131 | 0.499 | 1.136 |
|  |  |  | (0.397 - 1.467) | (0.586 - 2.183) | (0.185 - 1.342) | (0.549 - 2.352) |
| *Other inpatient ward* |  |  | 0.664 | 0.652 | 0.681 | 0.391 |
|  |  |  | (0.331 - 1.333) | (0.210 - 2.023) | (0.358 - 1.298) | (0.131 - 1.168) |
| *Surgery* |  |  | 4.050** | 4.745*** | 1.775 | 3.918*** |
|  |  |  | (1.079 - 15.21) | (1.889 - 11.92) | (0.370 - 8.503) | (1.490 - 10.30) |
| **Any inpatient days** (yes) |  |  | 0.229*** | 1.742 | 0.411*** | 2.038 |
|  |  |  | (0.130 - 0.405) | (0.520 - 5.837) | (0.233 - 0.722) | (0.660 - 6.291) |
| **Travel time to facility** (10-minute units) |  |  |  |  | 0.997 | 1.015 |
|  |  |  |  |  | (0.963 - 1.032) | (0.986 - 1.045) |
| **Time spent with HCW** (10-minute units) |  |  |  |  | 0.972 | 1.052 |
|  |  |  |  |  | (0.865 - 1.092) | (0.957 - 1.157) |
| **Accessed required medication** *(Ref – Yes)*  *No* |  |  |  |  | 21.87*** | 0.132*** |
|  |  |  |  |  | (11.42 - 41.86) | (0.0593 - 0.296) |
|  |  |  |  |  | 4.201*** | 0.910 |
| *None prescribed* |  |  |  |  | (1.944 - 9.079) | (0.558 - 1.481) |
| **Any fees paid to the facility** (Yes) |  |  |  |  | 0.492*** | 0.527 |
|  |  |  |  |  | (0.337 - 0.719) | (0.241 - 1.151) |
| Constant | 0.136*** | 1.850 | 0.342* | 0.920 | 0.371 | 0.667 |
|  | (0.0513 - 0.358) | (0.706 - 4.851) | (0.115 - 1.012) | (0.192 - 4.407) | (0.0710 - 1.938) | (0.107 - 4.172) |
| Observations | 4,052 | 4,052 | 4,052 | 4,052 | 3,588 | 3,588 |
| CI in parentheses. *** p<0.01, ** p<0.05 | | | | | | |
