## Supplementary File 2 - Survey questions for "Healthcare service user-reported quality of care in Malawi: a national multi-facility cross-sectional study"

### TLM Data Collection Tool 2: Patient Survey

#### Introduction

This tool covers patient characteristics including age, sex, reason for attendance, highest level of education, and household assets (to calculate wealth index), persistence in care seeking, costs and opportunity costs of attending facility, perceptions of quality of care, and health outcomes at two-week follow-up.

#### Patient details

RA Name *

- ○ [redacted]
- ○ [redacted]
- ○ [redacted]
- ○ [redacted]
- ○ [redacted]
- ○ [redacted]
- ○ [redacted]
- ○ [redacted]
- ○ [redacted]
- ○ [redacted]
- ○ [redacted]
- ○ [redacted]
- ○ [redacted]
- ○ [redacted]
- ○ [redacted]

Health Facility *

- ○ St Johns Mission Hospital
- ○ Mlambe Hospital
- ○ Zingwangwa Urban Health Centre
- ○ Malabada Health Centre
- ○ Queen Elizabeth Central Hospital
- ○ Chiradzulu District Hospital
- ○ Mua Mission Hospital
- ○ Kaporo Rural Hospital
- ○ Likoma/St Peters
- ○ Bwaila Hospital
- ○ Daeyang Luke Hospital
- ○ Dzenza Health Centre
- ○ Chiwe Health Centre
- ○ Lilongwe Central Hospital
- ○ Kabudula Community Hospital
- ○ Area 25 Health Centre
- ○ Mzuzu Urban Health Centre
- ○ Mzuzu Central Hospital
- ○ Lisungwi Health Centre
- ○ Nkhata Bay District Hospital
- ○ Chintheche Rural Hospital
- ○ Trinity - Fatima Hospital
- ○ Mlolo Dispensary
- ○ Bilira Health Centre
- ○ Sharpe Valley Health Centre
- ○ Bolero Rural Hospital
- ○ Mhuju Rural Hospital
- ○ Makwasa Estate Clinic
- ○ Pirimiti Health Centre
- ○ Zomba Central Hospital
- ○ Zomba Mental Hospital
- ○ Mianga Clinic

Name of respondent *

Provide the Age of Patient (years and months)

Age (Years) *

Age (Months) *

Sex of patient *

- ◯ Male
- ◯ Female

Date *

yyyy-mm-dd hh:mm

Reason for attendance *

- ◯ unwell
- ◯ accompanying some-one who is unwell
- ◯ for a test
- ◯ to receive test results
- ◯ for medicines
- ◯ other

If other, please specify *

Referred here from another facility? *

- ◯ Yes
- ◯ No

Highest education level of respondent *

- ◯ None
- ◯ Some primary
- ◯ Primary completed
- ◯ Some secondary
- ◯ Secondary completed
- ◯ Tertiary

#### Time and motion study involvement

Have you been given one or more paper numbers from my colleague after a consultation? *

*If yes -> ask to see the ticket(s) and record the number(s)*

- Yes
- No

Patient ID number 1 (from paper given to patient)

Patient ID number 2 (from paper given to patient)

#### Facility Visit

Date of arrival

*If the patient arrived before today, please ask about the date of arrival*

yyyy-mm-dd

Time arrived at facility *

hh:mm

Time left facility *

hh:mm

Time left facility cannot be earlier than time of arrival

Clinic *

- Outpatients - children
- Outpatients - adults
- Outpatients - general
- HIV clinic
- TB clinic
- Malaria clinic
- Cervical cancer
- Antenatal care clinic
- Antenatal care ward
- OBGYN clinic
- OBGYN ward
- Intrapartum and immediate newborn care
- Postnatal ward
- Postnatal care contact (maternal)
- Postnatal care contact (neonatal)
- Post abortion care
- Ectopic Pregnancy case management
- NCD clinics/inpatient
- Intensive and emergency care and recovery room (adult/paediatric)
- Surgical Clinic
- Surgical Ward
- Hypertension clinic
- Epilepsy clinic
- Maternity (labour and delivery) ward
- Neonatal ward
- Paediatric (children) ward
- Male ward
- Female ward
- Neonatal intensive care (NICU)
- Paediatric intensive care (PICU)
- High Dependency / Intensive Care Unit
- General emergency unit
- Paediatric emergency unit
- Diabetes clinic
- Surgery (operating theatre)
- Dental Clinic
- Eye Clinic
- Pharmacy
- Laboratory
- Radiology
- Dermatology
- STI Clinic
- Oncology Clinic
- Physiotherapy Clinic

Condition *

- HIV
- TB
- Mental Health
- Diabetes
- Hypertension
- Epilepsy
- Asthma
- Other NCD
- Screening Pregnancy
- ANC
- Facility delivery
- Postnatal Care
- Family Planning
- STIs
- Malnutrition (under five)
- IMCI
- Immunisation (under five)
- Malaria
- Pneumonia / ALRI
- Diarrhoea
- Other

If other, please specify *

#### » Time Spent With Health Workers

1

Health worker 1 *

*Who did you see today?*

- ◯ Doctors (medical officer)
- ◯ Clinical officers
- ◯ Clinical technicians
- ◯ Medical Assistants
- ◯ Nurses (state registered)
- ◯ Nurse midwife technicians
- ◯ Lab Officer
- ◯ Lab Technicians
- ◯ Lab Assistant
- ◯ Pharmacist
- ◯ Pharm Technician
- ◯ Pharm Assistant
- ◯ HSA (DCSA)
- ◯ Dental Officer
- ◯ Dental Therapist
- ◯ Dental Assistant
- ◯ Mental Health Staff (Clinician/Nurse)
- ◯ Nutrition Staff
- ◯ Radiographer
- ◯ Radiography Technician
- ◯ Sonographer
- ◯ Radiotherapy Technician
- ◯ Other

If other, please specify or comment *

*If not sure describe*

..................................................

How many minutes did you spend with the health worker? *

..................................................

Did you feel that you received appropriate care during your visit? *

- ◯ Yes
- ◯ No

Was a consultation conducted with you during your Visit? *

- ◯ Yes
- ◯ No

Was a previously undiagnosed condition diagnosed at this visit? *

- ◯ Yes
- ◯ No

Were you given a physical examination during your visit? *

- ◯ Yes
- ◯ No

Were you required to undergo any of the following tests as part of your visit? *

*Tick the options that apply*

- ☐ Blood test
- ☐ Urine Test
- ☐ X-ray
- ☐ MRI
- ☐ CT Scan
- ☐ Ultrasound
- ☐ None
- ☐ Other

If other, please specify *

........................................

Did you receive any specific treatment or prescribed medication during your visit? *

- Yes
- No

How can you rate the severity of your illness *

- Healthy
- Mild symptoms
- Moderate symptoms
- Severe symptoms
- Life threatening condition

#### Patients' access to medicines

During your visit today, were you able to access the medication prescribed to you at the health facility? *

- Yes
- No
- NA (I was not prescribed any medication)

(If answer to 1 is No) How do you plan to access these medications? *

- Come back to this health facility on another day
- Go to a different government health facility/pharmacy
- Go to a different private health facility/pharmacy
- Other

If other, please specify *

#### Patients' persistence in care-seeking

Have you sought care for this episode of this condition before?

- Yes
- No

If yes, where?

- At this health facility
- At a different health facility
- At a pharmacy
- In the community

Why are you seeking health care again?

- Did not receive any care before because the relevant healthcare worker was not present at the facility
- Did not receive any care before because the relevant healthcare worker was too busy
- Did receive care but not satisfied with it
- New instance of this condition that requires care
- Medication refill

#### Costs and opportunity costs of attending facility

For how many days have you stayed at the facility?

*If the patient arrived at the facility earlier than today (inpatient)*

How many minutes does it take you to get to this health facility?

Transport cost (one way) (MWK)

Food cost (MWK)

Accommodation cost (MWK)

Any payment made to facility (MWK)

What was your primary mode of transportation today? (COMING TO THE FACILITY)

- By foot (Wapansi)
- Bicycle (Njinga yakapalasa)
- Private car (Galimoto la munthu)
- Public car/bus including cab (Basi kapena taxi)
- Private motorcycle (Njinga yamoto yamunthu)
- Kabaza
- Other (Specify)

If other, please specify

What activities did you not do today as a result of you attending this facility?

How many people have accompanied you to the facility today?

Were you prioritised during triage?

- ◯ Yes
- ◯ No

#### Perceptions of quality of care (choose one for each question)

| New Question | Perception | Comment (probe if below good) |
|---|---|---|
| **Health worker consultation** | ◯ Very Bad ◯ Bad ◯ Neither bad or good ◯ Good ◯ Very good | |
| **Treated with respect** | ◯ Very Bad ◯ Bad ◯ Neither bad or good ◯ Good ◯ Very good | |
| **Waiting time** | ◯ Very Bad ◯ Bad ◯ Neither bad or good ◯ Good ◯ Very good | |
| **Treatment available** | ◯ Very Bad ◯ Bad ◯ Neither bad or good ◯ Good ◯ Very good | |
| **Overall** | ◯ Very Bad ◯ Bad ◯ Neither bad or good ◯ Good ◯ Very good | |

#### Household Assets

Do you have the following household assets

Electricity *

- ◯ Yes
- ◯ No

Radio *

- ◯ Yes
- ◯ No

Television *

- ◯ Yes
- ◯ No

Mobile phone *

- ◯ Yes
- ◯ No

Non-mobile telephone *

- ◯ Yes
- ◯ No

Computer *

- ◯ Yes
- ◯ No

refrigerator *

- ◯ Yes
- ◯ No

Koloboyi *

- ◯ Yes
- ◯ No

Paraffin lamp *

- ◯ Yes
- ◯ No

Torch *

- Yes
- No

Bed with mattress *

- Yes
- No

Sofa set *

- Yes
- No

ownership of agricultural land *

- Yes
- No

Ownership of farm animals *

- Yes
- No

Bicycle *

- Yes
- No

Motorcycle/scooter *

- Yes
- No

Car/truck *

- Yes
- No

#### Contact Details for Follow up

Is there a phone that we can use to call you in two to follow up on our conversation?

- Yes
- No phone

Phone Number for, 2 week follow-up

Next of kin

Phone Number for, 2 week follow-up
